# Inferring epidemiological parameters under an infectious phylogeography model with visitor dynamics

**DOI:** 10.64898/2025.12.16.25342415

**Authors:** Albert C. Soewongsono, Ammon Thompson, Michael J. Landis

## Abstract

During an outbreak, infectious disease can spread among populations through host movement, potentially fueling local outbreaks with their own epidemiological dynamics. However, it is difficult to know how often infections between populations are transmitted by diseased travelers infecting healthy residents when abroad, rather than by diseased residents infecting healthy travelers, who later return home with the new pathogen. In this paper, we introduce a phylogeographic model where pathogens spread through visitor dynamics, whereby hosts “visit” other populations for short trips before returning home. To do so, we used the stationary properties of an epidemiological compartment model with visitor dynamics to construct an approximation that is statistically accurate and computationally tractable for phylogenetic modeling. We applied our model to empirical infection data and travel statistics from the European SARS-CoV-2 pandemic. Inference under our model suggests that, in the early stages of the outbreak, SARS CoV-2 was more often “pulled” into new countries by returning travelers than “pushed” into new countries by visitors from the source country. Estimates of host movement–related parameter values under our visitor model suggest that competing migration models, with trips of indefinite length, may underestimate the magnitude of outbreaks caused by visitors. This study emphasizes the importance of carefully incorporating host movement dynamics into such models.

**Significance Statement:** Viral infections can spread among distinct locations by human travel. Being able to identify the source of a local outbreak – whether triggered by infected visitors or individuals returning home – is crucial. Here, we designed a phylogenetic model for modeling infectious disease transmission in spatially structured populations, explicitly accounting for realistic host movement. By accounting for “short trips”, our model can help inform travelrelated strategies for containing outbreaks.

---

**M**athematical models are crucial for understanding the dynamics of disease spread through populations which can help guide intervention strategies to contain the disease (1– 5). In mathematical epidemiology, researchers often model disease transmission dynamics by grouping individuals into different compartments based on their infection status and describe the time evolution of compartment sizes with coupled differential equations (6–8). For instance, the well-known SIR model describes the rate of change of the number of individuals among susceptible (S), infectious (I) and recovered (R) compartments (8). SIR models, and other models like it, are made possible by assuming such compartments interact in a well-mixed population, meaning each susceptible individual has an equal probability of being infected by any infectious individual (7).

In reality, many populations are spatially, ecologically, or behaviorally structured (7, 9, 10). This is concerning, as numerous theoretical and empirical studies have found assuming a population is well-mixed while ignoring population structure can yield misleading model estimates (11–13). To add more realism to compartmental models, researchers have designed metapopulation models where subpopulations are well-mixed, but transmission between subpopulations occur at a lower rate (12, 13). Citron et al. (14) examined two general models of transmission between subpopulations: the Flux model and the Simple Trip model (13). These two models differ in how disease jumps from one subpopulation to another. The Flux model treats host movements as indefinite migrations, where hosts change their place of residence and thus jumps are influenced by a single migration rate. Under the Simple Trip model, on the other hand, each host has a fixed place of residence, but randomly visits other locations for short periods before returning home. This creates brief windows of opportunity for an infectious visitor to transmit disease to the new location, or for a healthy visitor to become infected when away before going home. The transmission rate between locations is thus influenced by a visit rate and a return rate. These two models can produce conflicting estimates in many scenarios (14).

Pathogen spread is also studied within a phylogenetic context, where tree branching patterns and divergence times among sampled pathogens provide vital historical context for how outbreaks progress (15–19). Phylogeographic models in particular have become instrumental for reconstructing the spread of disease (20–23). These tools allow joint inference of model parameters and disease transmission history including the location and timing of the outbreak’s origin (24–26). Kühnert et al. (27), for instance, developed a probabilistic framework that allows for the joint reconstruction of phylogenies and epidemiological parameters, where the movement of infected hosts allow their pathogens to jump into new populations. Phylogenetic migration models of this kind are similar to the Flux model of Citron et al. (14) in that hosts (and their pathogens) travel itinerantly among locations.

Innovative phylogenetic migration models have been developed to associate pathogen movement with various kinds of travel data, including population flow between cities (28–31), individual flight history data (32), and international flight traffic (33, 34). However, phylogenetic models that only model indefinite migrations, rather than short trips, cannot make full use of publicly available travel data, such as the average number and length of trips taken between different countries. Models with more realistic mechanisms for host movements, informed by such data, could allow epidemiologists to quantify the relative importance that visitors and residents play in the spread of pathogens during an outbreak.

Here, we present a phylogeographic model that accounts for visitor dynamics to infer epidemiological parameters, host movement-related parameters, and disease transmission history from a serially sampled phylogenetic tree within an SIR framework. After introducing key mathematical details for the Visitor SIR model, we validate the model through simulation experiments, and then apply it to study transmission and movement dynamics for the SARS-CoV-2 outbreak in Europe (22). We compare the inference results under our model against alternative phylogenetic migration models and discuss how their differences relate to policy decisions during an outbreak.

## Model definition

We designed a phylogeographic model, called the Visitor SIR model, that allows disease to spread among populations in different locations through visitor dynamics (Fig. 1(a)). The Visitor SIR assumes that hosts remain at home (as residents) except during occasional trips to other locations (as visitors) before returning home again (13, 14). This leads to four distinct transmission modes: (1) resident-infects-resident, (2) visitor-infects-resident, (3) resident-infects-visitor, and (4) visitor-infects-visitor (Fig. 1(b)-(e)). To model these transmission modes, the Visitor SIR model uses a pair of states to represent the home and away locations for each individual: state (*i, j*) represents an individual from home location *i* that is currently visiting location *j*. Following suit, compartments for susceptible (S) and infected (I) hosts are subdivided by distinct home-away location pairs. That is, we have *S* = ∑_*i*_∑_j_ *S*_*ij*_ and *I* = ∑_*i*_∑_*j*_ *I*_*ij*_. Both susceptible and infectious individuals depart from home to visit new locations with the location-index rate from ***v*** = {*v*_*i* → *j*_ *}*, and return home with the appropriate rate from ***r*** = {*r*_*j* → *i*_ *}*. At equilibrium, some fraction of the population in each location is away from home, visiting another location, at any given time.

**Fig. 1.**
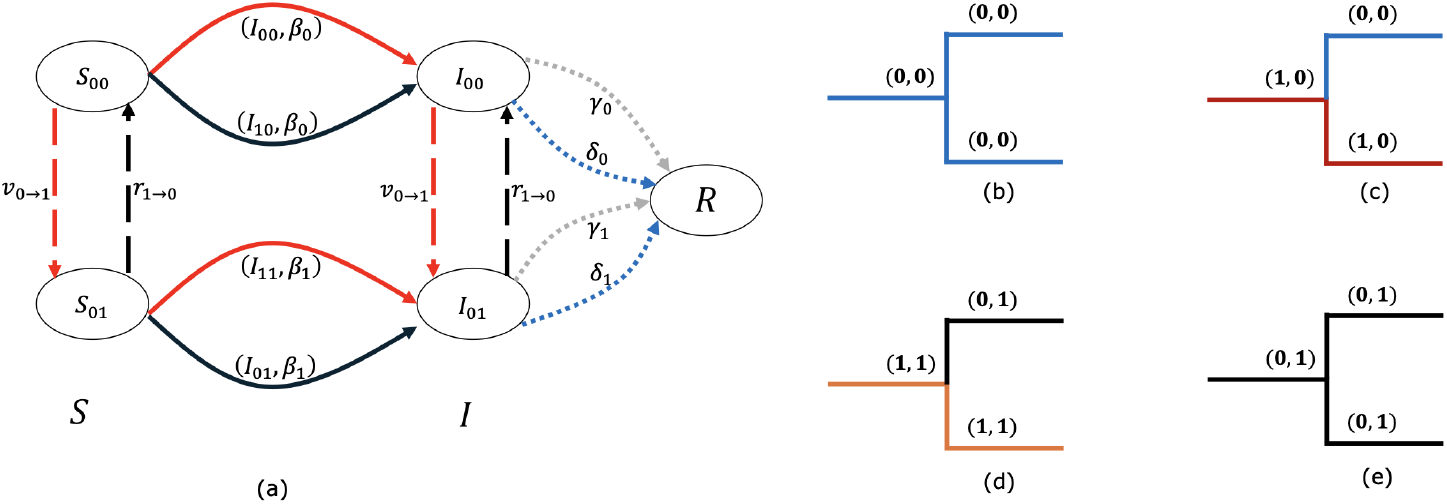
The full Visitor SIR model. (a) Transmission diagram under the full model, viewed from the perspective of a susceptible individual with home location in 0. By symmetry, the transmission dynamics for a susceptible individual with home location in 1 follows in the same manner. Compartments are represented by ellipses: *S*_*ij*_ for susceptible, *I*_*ij*_ for infectious, with home location *i* and current location *j*. Finally, *R* denotes recovered individuals irrespective of location. Arrows represent epidemiological transitions of hosts from one compartment (tail) into a new one (head) and are labeled with the corresponding rate parameters. Dashed arrows represent host movement events, either away from the home location through a depart event (red) with rate *v*, or back into the home location through a return event (black) with rate *r*. Solid arrows represent infection events by an infectious resident (red) or an infectious visitor (black), where the tuples indicate which infectious compartment caused the infection and the corresponding per-capita infection rate. For example, the top arcs involving *S*_00_ and *I*_00_ compartments indicate: (1) a resident-infects-resident event in location 0 with rate *β*_0_, *S*_00_ + *I*_00_ → 2*I*_00_; (2) a visitor-infects-resident event in location 0 with rate *β*_0_, *S*_00_ + *I*_10_ → *I*_00_ + *I*_10_. Dotted arrows represent removal events in location *j*, either by host recovery from infection (gray) with rate *γ*_0_ or by a sampling event with rate δ_0_. The other figures (b - e) illustrate four different cladogenetic event patterns under the Visitor SIR model, namely (b) resident-infects-resident, (c) visitor-infects-resident, (d) resident-infects-visitor, and (e) visitor-infects-visitor. A similar diagram for the approximate Visitor SIR model is shown in Supp. Fig. S1.

Branching events under the Visitor SIR model result from the new infection of a previously susceptible individual. An infectious individual (the infector) with home location *h* infects a susceptible with home location *i* in their shared, current location *j* at the per-capita infection rate of *β*_*j*_. We define ***β*** = {*β*_*j*_*}* in terms of standard epidemiological parameters, including the basic reproduction number 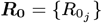, the percapita sampling rates **δ** = {δ_*j*_ *}*, and the per-capita recovery rates ***γ*** = {*γ*_*j*_ *}*, where 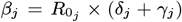. The location specific parameters 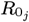, δ_*j*_, *γ*_*j*_ and *β*_*j*_ are defined for each current location *j* where the infection occurs rather than the home location of the infectious individual. Consequently, under the Visitor SIR model, the per-capita infection rate involving individuals *from* home-locations *h* and *i* who meet *in* current-location *j* is 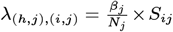 (see *SI Appendix* for derivation; Table S1 lists and summarizes all model variables).

Infections under the Visitor SIR model represent the four distinct modes of transmission, as described above. Residentinfects-resident events occur at rate *λ*_(*j,j*),(*j,j*)_ and are the only event type to represent a transmission scenario that remains contained within one location (“within-location infections”). Conversely, infections that eventually lead to transmission between two locations (“between-location infections”) are represented by three kinds of events, each of which involves at least one susceptible or infectious visitor: resident-infectsvisitor events occur at rate *λ*_(*j,j*),(*i,j*)_, visitor-infects-resident events occur at rate *λ*_(*i,j*),(*j,j*)_, and visitor-infects-visitor events occur at rate *λ*_(*i,j*),(*k,j*)_ where *i, k* ≠ *j* (see the *SI Appendix* for illustration). A migration model, in contrast, is not designed to distinguish among the three modes of between-location infection.

Unfortunately, an exact representation of the phylogenetic model likelihood under Visitor SIR dynamics is not known to us and appears intractable, since it requires marginalizing over the (unobserved) historical movements of susceptible and infectious hosts. We can avoid the need to track host movement among locations by assuming the proportion of susceptible individuals from location *i* visiting location *j* is approximately constant. To obtain this constant, we used the depart (***v***) and return (***r***) rates describing host movement to obtain the stationary distribution matrix, ***P***, of current locations for hosts. We then approximated the differential equation for changes in numbers of infected individuals under the full model, 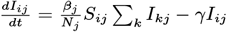, with the approximation, 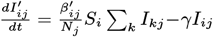, where 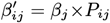 is the adjusted per-capita infection rate, and *S*_*i*_ = ∑_*j*_*S*_*ij*_ is the number of susceptibles with home location *i*. As a result, the rate at which an infector with home location *h* infects a susceptible with home location *i* while they are both in location *j* is 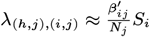 (see *SI Appendix* for the derivation). The definition of this rate has two important properties. First, the approximated rate is defined in terms of the home location, *i*, and current location, *j*, of the susceptible who is infected, but not in terms of the home location, *h*, of the infector; it does not matter where the infector is from, only that the infector shares a location with the susceptible. Second, the rate is a function of the local infection rate, *β*_*j*_, and the system of visitor depart (***v***) and return (***r***) rates, which directly links host movement to spatial infection outcomes. We then construct a tractable phylogenetic model likelihood in a manner analogous to Kühnert et al. (27), substituting in our state space and event rates (see *SI Appendix* for full description).

### Model validation

We compared the accuracy between the full and approximating versions of the Visitor SIR model in a three-location system. To do so, we used MASTER (35) to simulate one phylogenetic tree using the same simulating conditions under both models, and then compared the number of infections over time across compartments *I*_*ij*_ (*Materials and Methods*). Both models produced the equivalent compartment sizes *I*_*ij*_ through time, indicating the mathematical approximation is accurate and that the simulation framework we later use to validate the inference method is correct (Figs. S2 − S4 in *SI Appendix*).

Next, we compared the performance of the Visitor SIR model against two Migration SIR model variants (27) in a phylogenetic context using RevBayes (36) with the Tensorphylo plugin (37). Unlike a Visitor SIR model, hosts under the two Migration SIR models have no home location and instead randomly move (migrate) between locations *i* and *j* at rate *m*_*i*→*j*_. For the Simple Migration SIR, host movement only occurs along branches (i.e., during anagenesis), not at branching events with infection (i.e., no cladogenesis). The only infection type under this model is a within-location infection which occurs at rate of *λ*_*i,i*_ in location *i*. Under a Cladogenetic Migration SIR model, infectious hosts in location *i* infect susceptible hosts in location *j* at rate *λ*_*i,j*_. Importantly, these between-location infection rates do not depend on the migration rate, meaning the model treats anagenetic and cladogenetic movement as categorically different kinds of host movement. Moreover, while infection between locations during cladogenesis implies movement of the susceptible and/or infectious individual, that movement is not explicitly represented in the Migration SIR model, making it difficult to determine if visitors tend to import or export more infections. The Supplement provides a complete description of both Migration SIR models we used for comparisons in our study.

### Visitor SIR model estimates are accurate

We measured how reliably the Visitor SIR model recovered true data-generating parameters through a simulation experiment (see *Materials and Methods*). Posterior mean estimates and 80% highest posterior density (HPD80) intervals demonstrate that our Visitor SIR model in RevBayes accurately inferred both epidemiological parameters, *R*_0_ and δ, the within-location infection rate, *λ*_(*j,j*),(*j,j*)_, and the between-location infection rate, *λ*_(*j,j*),(*i,j*)_, when the simulating model and the inference model (Visitor SIR) matched (Fig. 2(a)). Accuracy remained high for other experiments that explored alternative numbers of locations and relationships between the visit depart and return rates (Figs. S5, S9 and S13 in the *SI Appendix*). While most parameters under most conditions produced accurate estimates with correct coverage (near 80%), the basic reproduction number, *R*_0_, and movement rates, *v* and *r*, occasionally fell outside the expected 95% confidence interval in a Bernoulli experiment with 116 samples (4 in 9 cases for *R*_0_ and 1 in 9 cases for movement rates) (see *SI Appendix*).

**Fig. 2.**
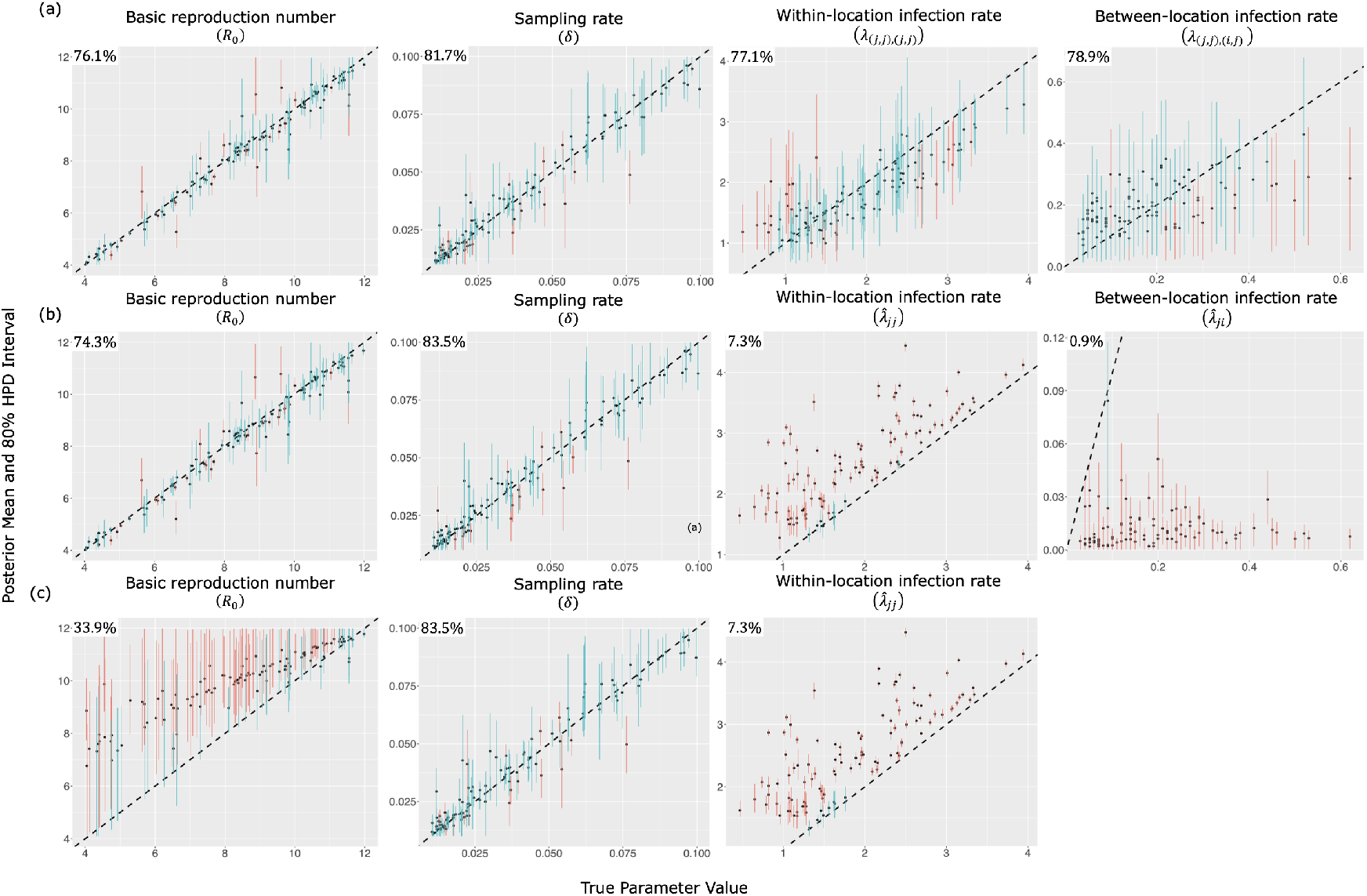
Quality of parameter estimation and coverage across the inference models, namely (a) the Visitor SIR, (b) Cladogenetic Migration SIR, and (c) Simple Migration SIR models where the simulation model (the true model) is simulated under the same Visitor SIR model with 5 locations as used for inference (see *SI Appendix*). Plots show true parameter values on the *x*−axis and the estimated values on the *y*−axis. For each plot, 80% HPD intervals, which covers the truth, are shown in blue, and 80% intervals, which do not cover the truth, are shown in red. The coverage percentages are shown in the top left of each plot.

### Migration SIR models misestimate infection rates

To characterize how misspecifying movement dynamics influences phylogeographic parameter inference, we measured the accuracy of the Simple Migration SIR and Cladogenetic Migration SIR models when fitted to data simulated under the Visitor SIR model. We found that the coverage for both *R*_0_ and δ under the Cladogenetic Migration SIR were satisfactory, while *R*_0_ under the Simple Migration SIR model tended to be increasingly overestimated as the true values of *R*_0_ decreased (Fig. 2). This is expected because the misspecified Simple Migration SIR model assumes all infected individuals in a location are residents while, in truth under the simulating model, some of them will be visitors that would then carry the disease elsewhere. For both models, accuracy and coverage for the within-location infection rate parameter (*λ*_*i,i*_) under both migration models was always high, caused by a constant overestimation bias of about 0.5 units. Accuracy and coverage for the between-location infection rate parameter (*λ*_*i,j*_) under the Cladogenetic Migration SIR model was also low, but it was instead typically underestimated to about 10% of the true value. Although it is difficult to interpret parameters values across different models with different state spaces, these estimates show that Visitor and Migration SIR models lead to similar estimates for both *R*_0_ and δ, but Migration SIR models tend to misestimate infection rates when data are generated through visitor dynamics.

### European SARS-CoV-2 outbreak

We analyzed a phylogenetic tree of the 2020 European SARS-CoV-2 outbreak produced by Nadeau et al. (22) with the Visitor SIR model and Cladogenetic Migration SIR model (see *Materials and Methods*). Both models estimated si milar numbers of secondary infections (*R*_0_) in France and Germany, but showed minor differences for Hubei, Italy and Other European countries. The Cladogenetic Migration SIR model predicted slightly higher *R*_0_ values in Hubei and Italy compared to the estimates from the Visitor SIR model, and slightly lower values for Other European countries (Fig. 3(a)). Under our Visitor and Cladogenetic Migration SIR models, France had the highest value of *R*_0_, followed in order by Other European nations, Italy, Germany, and Hubei. Posterior mean values of *R*_0_ for each location under the Visitor SIR model also fell within the per-location 95% HPD intervals previously reported by Nadeau et al., who used a Simple Migration model (no pathogen movement during cladogenesis; their Fig. S2) with unique migration rate parameter for each pair of locations and a single non-empirical prior describing those rates. In terms of ranking, our result slightly differed from Nadeau et al. (22), swapping the order for Italy and Other European nations.

**Fig. 3.**
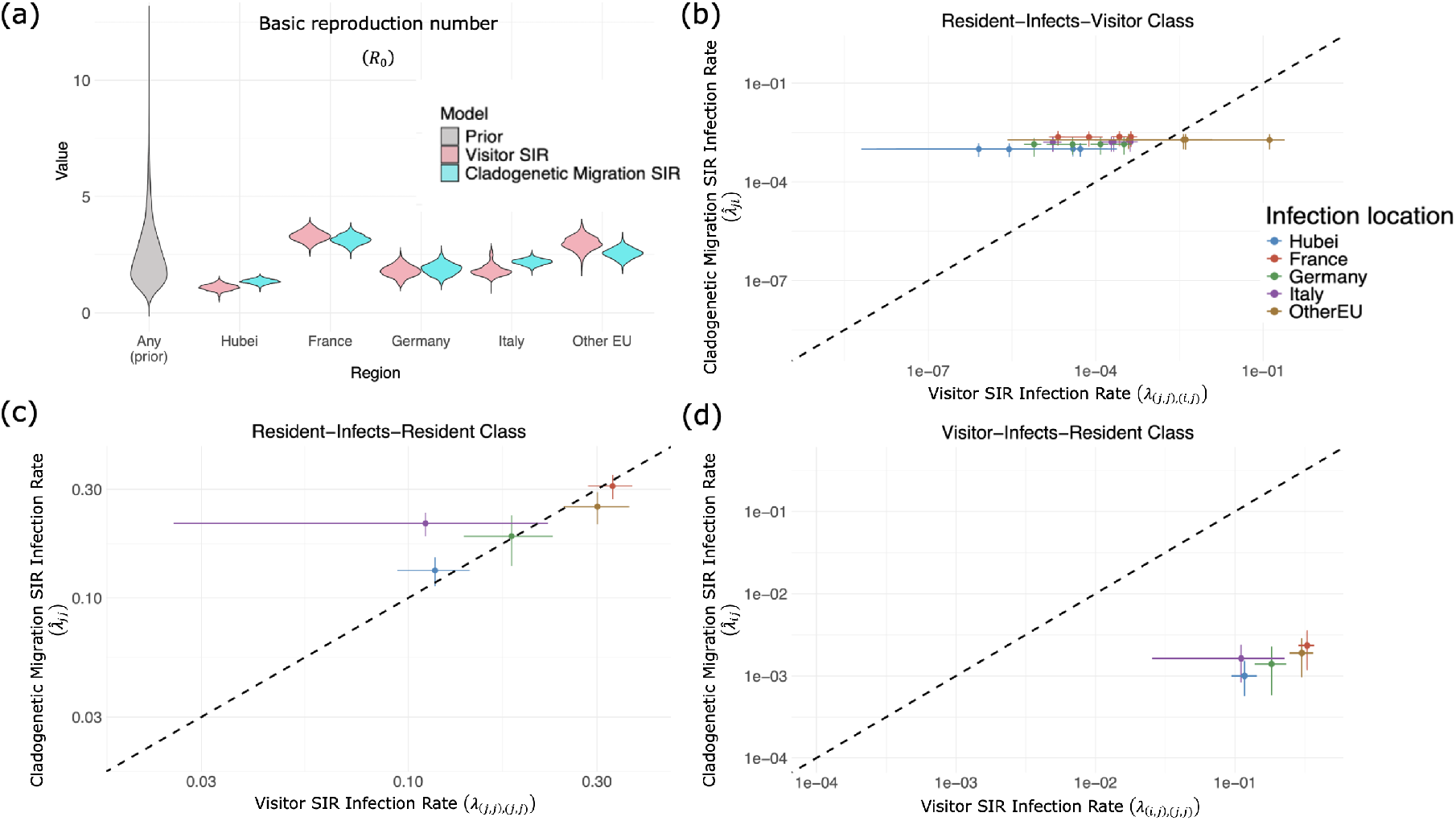
(a) Estimated basic reproduction number (*R*_0_) across different locations between Visitor SIR (pink violin plots) and Cladogenetic Migration SIR (cyan violin plots) models, along with prior distribution (gray violin plot). Note that we use the same prior distribution for all locations (“Any (prior)”). We used the empirical phylogenetic tree derived in (22). The 95% highest posterior density (HPD) for each location is, as follows: [0.87, 1.37] (resp. [1.17, 1.55] for Cladogenetic Migration SIR) for Hubei; [2.83, 3.74] (resp. [2.69, 3.50] for Cladogenetic Migration SIR) for France; [1.31, 2.32] (resp. [1.40, 2.38] for Cladogenetic Migration SIR) for Germany; [1.45, 2.56] (resp. [1.91, 2.41] for Cladogenetic Migration SIR) for Italy; [2.34, 3.51] (resp. [2.09, 2.94] for Cladogenetic Migration SIR) for Other European countries. (b)-(d) Infection rates under fitted Visitor SIR (*x* axis) and fitted Cladogenetic Migration models (*y* axis) given the empirical phylogenetic tree from Nadeau et al. (22) and priors setup on Table 1. Each colored dot represent posterior mean of the corresponding rate from both models and the lines through each dot represent the 95% HPDs from both models. Within the “Resident-Infects-Resident” class, we compare within-location infection events in the migration model and resident-infects-resident events in the visitor model. Within the “Visitor-Infects-Resident” class (resp.”Resident-Infects-Visitor”), we compare between-location infection events in the migration model and visitor-infects-resident (resp. resident-infects-visitor) events in the visitor model.

Next, we compared the infection rates within and between locations for our two models. The Cladogenetic Migration SIR model inferred within-location infection rates (*λ*_*j,j*_) that were similar to the equivalent resident-infects-resident event rates (*λ*_(*j,j*),(*j,j*)_) estimated under our Visitor SIR model (Fig. 3(b)).

**Table 1.** Values and priors for the parameters from different models used for the empirical analysis. Our study considered three models: Visitor SIR and Cladogenetic Migration SIR models and Simple Migration models as described in this paper. For other parameters besides the host movement parameters in both models, we used the same prior distribution and values for the LDBDS model as in Nadeau et al. (22). *σ*_*i*_ is the standard deviation of the lognormal distribution for location *i, σ* = {0.8, 0.1, 0.1, 0.1, 0.8*}*

| Notation | Visitor SIR | Cladogenetic Migration SIR | Nadeau's LDBDS | Description |
| --- | --- | --- | --- | --- |
|  | Priors | Priors | Priors |  |
| $R_0$ | Lognormal(0.8, 0.5<sup>2</sup>) | Lognormal(0.8, 0.5<sup>2</sup>) | Lognormal(0.8, 0.5<sup>2</sup>) | basic reproduction number across all locations |
| $\gamma$ | 0.1 | 0.1 | 0.1 | fixed recovery rate (days<sup>-1</sup>) |
| $s_0$ | U(0,0.15) | U(0,0.15) | U(0,0.15) | sampling proportion in Hubei |
| $s_1$ | U(0,0.093) | U(0,0.0.093) | U(0,0.093) | sampling proportion in France |
| $s_2$ | U(0,0.10) | U(0,0.10) | U(0,0.10) | sampling proportion in Germany |
| $s_3$ | U(0,0.005) | U(0,0.005) | U(0,0.005) | sampling proportion in Italy |
| $s_4$ | U(0,0.057) | U(0,0.057) | U(0,0.057) | sampling proportion in Other European nations |
| $v_{i \rightarrow j}, \forall i, j$ | Lognormal($\ln(v_{i \rightarrow j}^e) - 0.5 \times \sigma_j^2, \sigma_j$) | — | — | per-capita depart rate from $i$ to $j$ |
| $r_{j \rightarrow i}, \forall i, j$ | Lognormal($\ln(r_{j \rightarrow i}^e) - 0.5 \times \sigma_i^2, \sigma_i$) | — | — | per-capita return rate to $i$ from $j$ |
| $m_{i \rightarrow j}, \forall i, j$ | — | Lognormal($\ln(v_{i \rightarrow j}^e) - 0.5 \times \sigma_j^2, \sigma_j$) days<sup>-1</sup> | Lognormal(0,1) year<sup>-1</sup> | migration rate from $i$ to $j$ |
| $p$ | — | U(0,1) | — | Cladogenetic probability of observing no change in state |

For both models, France and Other European countries exhibited the highest within-location infection rates, while Hubei had the lowest rate among all the other locations. This finding aligns with our *R*_0_ estimates under both models, where France and Other European countries have the highest infections, and Hubei has the lowest (Fig. 3(a)). When comparing the visitor-infects-resident event rates for the Visitor SIR model (*λ*_(*i,j*),(*j,j*)_) against the between-location infection rate for the Cladogenetic Migration SIR model (*λ*_*i,j*_), the Visitor SIR model estimated higher rates (Fig. 3(d)). In particular, France and Other European countries are the two locations where a visitor from other locations is most likely to infect a resident in these two locations. In contrast, when comparing between-location infection rates from the Cladogenetic Migration SIR model (*λ*_*i,j*_, as above) to the resident-infects-visitor event rates under the Visitor SIR model (*λ*_(*j,j*),(*i,j*)_), the Visitor SIR model almost always infers lower transmission rates (Fig. 3(b)).

To further examine the role that visitors and residents played in fueling the European outbreak, we visualized a single history of ancestral states on the empirical tree under our model (Fig. 4(a)). Most infection events leading to sampled lineages (e.g., branching events) involved infectious residents infecting other residents. These are followed in frequency by infection events resulting from host movement, involving either susceptible or infectious individuals. Additionally, we observe host movements along branches that occur after infection events. However, it is difficult to determine whether these movements are directly associated with infection events, as unsampled sister lineages do not appear in the phylogeny.

**Fig. 4.**
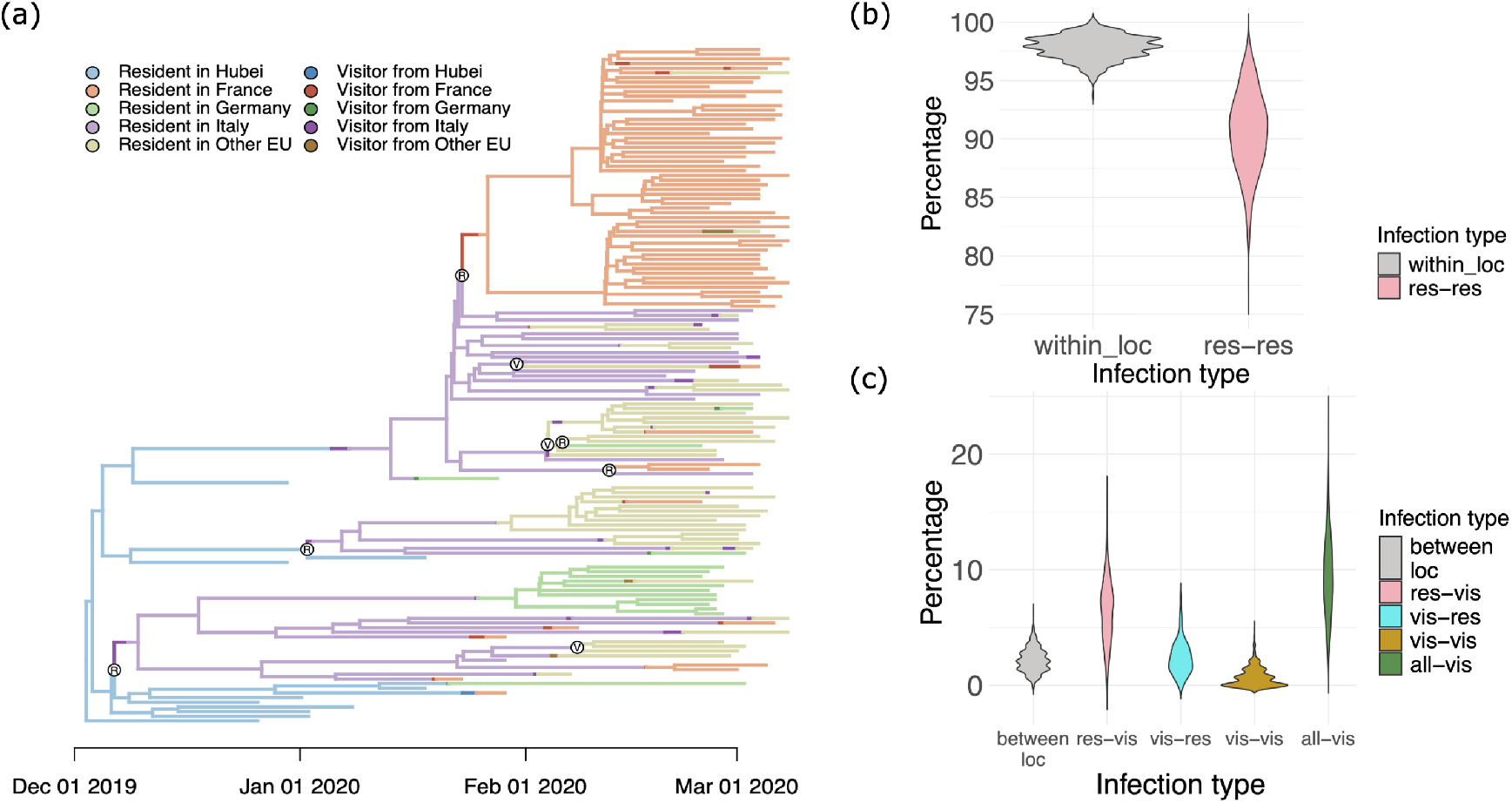
(a) One stochastically mapped state history under our Visitor SIR model fitted to empirical data from Nadeau et al. (22). Possible locations are Hubei, France, Germany, Italy, and Other European nations (Other EU). Lighter colors represent infectious residents in different locations and darker colors represent visitors currently visiting different locations. The internal node labels indicate the source of infection, whether it came from a visitor (labeled “V”) or from a resident (labeled “R”). The internal nodes that correspond to resident-infects-resident infection events are not labeled. (b)-(c) Summary of all possible lineage histories from stochastic mapping under fitted Visitor SIR model and fitted Cladogenetic Migration SIR model. The top panel shows the percentage of events that are within-location infection events under the migration model (“within_loc”, mean = 97.835%) and the percentage of events that are resident-infects-resident events under the visitor model (“res-res”, mean = 90.598%). The bottom panel shows the percentage of events that are between-location infection events under the migration model (“between_loc”, mean = 2.165%) and the percentage of events that are resident-infects-visitor (“res-vis”, mean = 6.353%), visitor-infects-resident (“vis-res”, mean = 2.360%) and visitor-infects-visitor (“vis-vis”, mean = 0.689%) under the visitor SIR model. We also plotted the distribution of infection events that involve at least one visitor under the visitor SIR model (“all-vis”, mean = = 9.402%).

The pattern of infection types portrayed in Figure 4(a) is preserved when looking at ancestral state histories across MCMC samples. That is, the resident-infects-resident infection is the most common infection type under our model, followed by resident-infects-visitor, visitor-infects-resident, and visitor-infects-visitor, respectively (Fig. 4(b)-(d)). This is not surprising because the most common infection type requires neither the infector nor the newly infected individual to move from their shared home location. Similarly, the within-location infection events were more common than the between-location infection events under the fitted Cladogenetic Migration SIR model, looking across all possible histories from stochastic mapping (Fig. 4(b)-(d)). Under the fitted Visitor SIR model, resident-infects-visitor events were more common than visitorinfects-resident events despite the inferred per capita rates showing the opposite pattern with visitor-infects-resident rates being greater than the resident-infects-visitor rates (Fig 3(b) and (d)). This apparent contradiction is because the number of infectious residents is much larger than the number of infectious travelers for most of an outbreak, resulting in a higher probability of infection spreading *to* visitors rather than *from* visitors. As expected, the infection type that requires movement from both individuals was the rarest (i.e., visitor-infects-visitor). A key insight provided by our model is that, during the earliest phase of the European outbreak, SARS-CoV-2 appears to more likely have been “pulled” into new locations by returning visitors rather than “pushed” into new locations by infectious travelers from source locations.

## Discussion

Our study introduces the Visitor SIR model, a phylogeographic model in which pathogens spread among structured populations when hosts visit locations away from home. Whereas standard migration-based models of phylogeography assume hosts randomly wander among different locations for indefinite periods of time (20, 27), hosts under the Visitor SIR model have a home they return to after each visit to another location. These two models are respectively analogous to the Flux and Simple Trip models described by Citron et al. (14). Under the Visitor SIR model, disease transmission between locations requires at least one individual to be visiting, or visiting from, the place of infection. This fundamentally links the between-location rate to underlying visitor depart and return dynamics that govern host movement, while also giving rise to three distinct between-location modes of disease transmission: resident-infects-visitor, visitor-infects-resident, and visitor-infects-visitor scenarios. In contrast, standard Cladogenetic Migration SIR models do not account for the short trips hosts make between home and away locations, treat anagenetic (along branches) and cladogenetic (during branching) migration events as unrelated phenomena, and cannot distinguish among three modes of between-location disease transmission.

Introducing more realism to how phylogeographic models represent host movement behavior carries some additional costs. First, our visitor dynamics require that individuals are associated with a home location and a current location. This squares the number of possible location-states used by a standard migration model, which results in slower likelihood evaluations, and it roughly doubles the number of movement parameters, which can complicate parameter inference. Neither of these issues prohibited our analysis involving five locations (i.e., 25 home-current locations), but this would scale poorly if the number of locations is large. Additionally, infection rates under visitor dynamics are functions of the distribution of individuals who are at home or traveling at a particular time. Because this exact headcount is generally unknown, we used the stationary probabilities induced by the visitor depart and return rates to approximate the typical abundance of local susceptibles. We found that simulations that assume realistic demographic conditions using either the full or approximating Visitor SIR models produced nearly identical disease prevalence curves. However, this must be verified for the unique conditions of each inference problem.

We used Bayesian coverage properties to assess the correctness of our Visitor SIR model and method implementation. We assessed this by analyzing simulated data, which shows our model can accurately recover the true parameter values used to generate each phylogeographic dataset. For simplicity, we assumed equal rates for movements between all pairs of location in both simulating and inference model for this experiment (see *Materials and Methods*). Epidemiological parameters for the basic reproduction number (*R*_0_) and sampling rate (δ) were estimated with high accuracy under scenarios with three, four, or five locations, and scenarios where the visit depart rates were less than, equal to, or greater than return rates (Fig. 2(a); Supp. Fig. S6; Supp. Fig. S10; and Supp. Fig. S14). Within-location and between-location infection rates were also inferred with acceptable accuracy, but reduced precision, when compared to the epidemiological parameters (Fig. 2(a) and Supp. Figs. S7-9, S11-13, S15-17). We were also able to infer the ratio of visit depart versus return rates, but with fairly low precision (Supp. Figs. S6, S10, S14); this motivated us to use empirical priors to inform these rates. About 1.8 of 36 (5%) cases examined (Supp. Figs. S6, S10, S14) should produce coverage levels that are outside the range of 72% to 87% by sheer chance. Coverage for δ, *λ*_(*jj,jj*)_, and *λ*_(*jj,ij*)_ are all acceptable, however in four instances coverage for *R*_0_ was lower than expected under the Visitor SIR model (Supp. Figs. S11-S13, S17). This may be explained by if some of our SIR-simulated trees passed the exponential phase of disease spread (i.e., model violation). Bayesian coverage levels indicated the simulator, inference method, and model design each operated as intended.

Across all simulations under the equal rates assumption for movement-related parameters, the Simple and Cladogenetic Migration SIR models accurately estimated sampling rates and overestimated within-location infection rates. The Cladogenetic Migration SIR model additionally estimated basic reproduction number accurately, but vastly underestimated between-location infection rates, while the Simple Migration SIR model, which has no between-location infection event type, overestimated the basic reproduction number. These results show that models which forbid between-location infections, such as the Simple Migration SIR model, will lead to biased estimates of the basic reproduction number. In addition, models that represent host movement among locations as indefinite migration events rather than short trips will inflate the within-location infection rate. Also, the Cladogenetic Migration SIR model will underestimate the between-location infection rate, potentially masking the true contribution of travelers to the spread of disease.

When applied to the initial European SARS-CoV-2 outbreak in early 2020, and allowing each pair of locations to have its own movement rates for both models, the Visitor SIR model largely agrees with the original study by Nadeau et al. (22) in estimating local *R*_0_ parameters. However, our model found that France had the highest number of secondary infections, followed by Other European countries, Italy, and Germany, in that order. This slightly differs from the previous finding in Nadeau et al. (22) in which they inferred that Italy has the second most infections, followed by Other European countries. Differences in these results may reflect differences in model design, as Nadeau et al. (22) used the equivalent of our Simple Migration model with unique migration parameters for each pair of locations, and applied non-empirical priors to their movement parameters.

Furthermore, the estimates for the *R*_0_ parameters are similar between our Visitor SIR and our Cladogenetic Migration SIR model for some countries, such as in France and Germany, but differ for other countries (Fig. 3(a)). Interestingly, our simulation experiment found that the Visitor SIR and Cladogenetic Migration SIR model produce highly similar *R*_0_ values when movement-related parameters are constrained to be equal for all locations (Fig. 2; Supp Figs. S6-S17). This discrepancy supports the notion that complex models are more capable of extracting subtle differences in location-specific parameters. As for infection events that involved at least one visitor, the Cladogenetic Migration SIR model estimated resident-infects-visitor (i.e., between-location infection) event rates that were often about 10 times faster (Fig. 3(b)), and visitor-infects-resident (i.e., between-location infection) event rates that were often 100 times slower, than those under the Visitor SIR model (Fig. 3(d)). The bias in estimation under the Cladogenetic Migration SIR model is consistent with results from our simulation-based experiments that assume equal movement rates across all location pairs. However, the direction of the bias differs, possibly due to the different priors used in simulation versus empirical analysis and differences in assumptions on movement parameters (Fig. 2; Supp Figs. S6–17).

Ancestral infection reconstruction of resident and visitor hosts among different locations generally supports the residentinfects-resident scenario as the most common mode of transmission (Fig. 4). Across all possible stochastic mappings of ancestral states under our fitted model, about 10% of branching events involved at least one visitor, with roughly twice as many residents infecting visitors as visitors infecting residents (Fig. 4(c)). In addition, infections repeatedly moved among locations along the phylogenetic branches, between observed branching events (i.e., during anagenetic host movement, or as part of a between-location infection event where one daughter lineage went unsampled). No branching events in the tree assigned the highest probability to the infection being associated with a visitor. However, each stochastically mapped tree in the posterior contained at least a few branching events that involved at least one visitor. Together, this means that it is certain that some branching events in the tree involved visitors, but it is difficult to know with any certainty exactly which branching events those are. It may be that by increasing the complexity of the host movement model through visitor dynamics, estimation bias is replaced with (correct) levels of uncertainty, as previously found by De Maio et al. (38) in their work on structured coalescent models for viral phylogeography.

We anticipate that performance for the Visitor SIR model can be improved further by adapting existing techniques developed for migration-based approaches and/or incorporating various types of travel data (39). For instance, using time-stratified visit depart and return rates under an “epoch” model (40) that was informed by travel time-series data would help quantify the role that visitors play on a seasonal basis (41), during holiday seasons (42), and before-versus-after travel restrictions (34). Allowing visit depart and return rates for infectious individuals, who might be reluctant to travel and potentially spread disease or wait longer to return if sick, to differ from those for (healthy) susceptible individuals could rebalance whether visitors or residents are more likely to be the infector for between-location transmission (43). Travel history information about the known or inferred home locations of sampled hosts, which we treated as ambiguous, could also help resolve which transmission events were associated with visitors rather than residents (32). Lastly, correlating the properties among locations with visit depart and return rates using generalized linear models, as introduced by Lemey et al. (20), could produce more realistic host travel patterns where, for example, long-distance trips tend to be less frequent but longer in duration.

Although our study focuses on the spatial movements of hosts among geographically structured populations, the Visitor SIR model is also appropriate for use with other structured populations. For example, in humans, individuals may temporarily enter social conditions that increase risk of infection (e.g., loss of reliable shelter (44)) before returning to more stable, low-risk conditions. In many species of other primates, individuals may primarily interact with others in their social group, but occasionally interact with individuals from other groups upon chance encounters, creating opportunities for intergroup, or even interspecific, pathogen transmission (45–48). Both cases create new opportunities to associate behavioral data and theory into the phylodynamic study of disease spread.

Phylogeographical models for studying epidemiology have grown increasingly realistic, both in terms of their mathematical construction and their ability to interface with a greater diversity of empirical data types. Our work introduces new phylogenetic modeling techniques to accommodate the temporary and intermittent movement of hosts between their home locations and the places they visit. We hope our study stimulates continued discussion about the complex relationship between host movement and pathogen spread.

## Materials and Methods

### Bayesian modeling framework

For all Bayesian analyses in this study, we used RevBayes (36) to construct our phylogenetic models and to generate samples from the posterior distribution using MCMC. In addition, we used the Tensorphylo plugin (37) for <monospace>RevBayes</monospace> to accelerate the computation of multitype birth-death likelihoods.

### Bayesian coverage experiment

We analyzed 116 transmission trees simulated using MASTER (35) for each simulation scenario using our Visitor SIR model. These scenarios differed in the number of locations (3, 4, and 5) and relationships between visit depart and visit return rates (*v*_*i*→*j*_ *< r*_*j*→*i*_, *v*_*i*→*j*_ = *r*_*j*→*i*_, and *v*_*i*→*j*_ *> r*_*j*→*i*_) (see *SI Appendix*). We assessed how reliably our model infers parameters when the inference and simulating models match. We used Revbayes and TensorPhylo for inference. MCMC analyses ran for 7, 500 generations with sampling every 10 iterations, producing effective sample sizes (ESS) of *>* 200 for all key parameters, and discarding the first 10% of the chain as burn-in. All inference jobs were run on Washington University in St. Louis’ RIS cluster. For 116 transmission trees with target coverage of 80% for each parameter, we expected the mean coverage would fall within the [72%, 87%] interval for 95% of tested parameters.

We assumed our data-generating parameters followed these distributions: *R*_0_ ∼ Unif[4, 12], *γ* ∼ Logunif[0.1, 1] (mean recovery time between 1 to 10 days; treated as clinically “known” and fixed during inference), δ ∼ Logunif[0.01, 0.1] (mean sampling time between 10 and 100 days). Both visit depart and visit return rates were drawn from either Logunif[0.01, 0.1] or Logunif[0.1, 1.0] for all pairs of locations *i* and *j*, depending on the assumed relationship (*<*, =, *or >*) between the two rates. Note that here we used a simplified Visitor SIR model where the depart rates (resp. the return rates) were constrained to be equal across each pair of home and away locations. In experiments where we added more locations, we multiplied the bounds for drawing the depart and return rates by 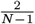 where *N* indicates the number of locations. This kept the total sum of rates of departing from location *i* into other locations and the total sum of rates of returning to location *i* from other locations constant in all experiments.

### Model comparison

Using simulated datasets from *Coverage experiment*, we examined how well the migration models perform, namely the Cladogenetic Migration and Simple Migration SIR, when fitted to data simulated under our more complex Visitor SIR model (see *SI Appendix*). To match the assumption on the movement-related parameters used in the simulating model, we also assumed that the migration rates were constrained to be equal across all pairs of locations. In both Migration SIR models, we generated Bayesian coverage plots for *R*_0_ and *γ* with at the 80% coverage level. Next, we defined a within-location infection event and a between-location infection event across these three models, and then compared their rates. Under the Visitor SIR, we defined a within-location infection as a resident-infects-resident event type (see Fig. S4(b) in *SI Appendix*) with rate given by *λ*_(*j,j*),(*j,j*)_. For the Migration SIR models, a within-location infection event was defined to occur in the current location and does not involve the movement of the newly infected individual (see Fig S4(a) in *SI Appendix*), with rate given by 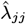. For this experiment, we then defined a between-location infection under the Visitor SIR model as a resident-infects-visitor infection, where the newly infected individual in the current location *j* returns to their home location in *i* (see Fig. S4(d) in *SI Appendix*), with rate given by 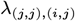. For the Cladogenetic Migration SIR model, it was defined as the infection of an individual in location *j* who then immediately moves into a new location *i* (see Fig S4(c) in *SI Appendix*), with rate given by 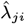.

### SARS-CoV-2 in Europe analysis

We applied our Visitor SIR model and Cladogenetic Migration SIR model to data from the phylodynamic study of the 2020 SARS-CoV-2 pandemic in Europe, which was originally assembled and analyzed by Nadeau et al. (22). Their study produced a phylogenetic tree with samples from five locations: three European countries (France, Germany and Italy), an aggregation of 16 Other European nations, and Hubei in China. To infer the tree, they used a Bayesian location-dependent birth-death-sampling (LDBDS) model with carefully constructed empirical priors. We used Revbayes and TensorPhylo for inference. Most parameters across these models obtained ESS values of *>* 100.

Our study used the main consensus tree of Nadeau et al. (their Fig. 1) for our analyses using Visitor SIR and Cladogenetic Migration SIR models. Moreover, since the pathogens are only associated with the (current) sample locations, and our model assumes each pathogen has a pair of home and current locations as states, we treated the unknown home locations for all individuals as ambiguous (see *SI Appendix* for the detailed transformation). Our empirical analysis used a Visitor SIR model where the visit depart and visit return rates can vary for each pair of home and away locations. By allowing the movement rates to vary between locations, we could demonstrate the flexibility of the model in using travel information. We therefore designed empirical priors for these host movement parameters using the 2019 tourist travel data (https://ec.europa.eu/eurostat/web/tourism/database) that contained information about the total number of trips and duration of trips between countries of interest in Europe and China. We rescaled the total number and duration of trips to and from China by the ratio of Hubei’s population to China’s population, in order to estimate the corresponding values for Hubei.

We approximated the daily, per-capita depart rate from home location *i* to an away location 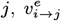 by taking the inverse of the expected number of trips from *i* to *j* per person per day, 𝔼 [#trips_*i*→*j*_] (see Eq. (1)).

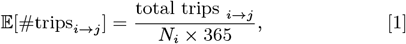

where total trips _*i*→*j*_ is the total number of trips between *i* and *j*, and *N*_*i*_ is the population size of country *i*. The population sizes of each country of interest are obtained from https://www.ons.gov.uk/. Furthermore, the daily, per-capita return rate from an away location *j* to home location *i*, 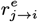, was approximated by taking the inverse of the expected duration of stay in *j* from *i* per trip, 𝔼 [#stays_*j*→*i*_] (see Eq. (2)).

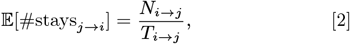

where *N*_*i*→*j*_ is the total number of nights spent in location *j* by visitors from location *i* per year, and *T*_*i*→*j*_ is the total number of trips to location *j* from location *i* per year. Note that some countries in the dataset had missing travel information for certain destinations (see Table S5 in *SI Appendix*). In such cases, we approximated the mean number of trips (and, respectively, the duration of stay) from those countries to a given location by rescaling the corresponding mean across all other countries to that location, using the population size of each country with missing data.

Finally, we used these empirical depart and return rates as the mean of lognormal distribution applied as a prior for these rates. For the Cladogenetic Migration model, we used the same prior used for the depart rates in the Visitor SIR model to describe movement parameters *m*_*i*→*j*_. For all other parameters, we used the same prior settings as used in Nadeau et al. (22). Table 1 shows the comparison between priors used in our study and priors used in Nadeau et al. (22).

To assess model performance, we compared the estimate for *R*_0_ and corresponding infection rates for all locations under the Visitor SIR and the Cladogenetic Migration SIR. Specifically, we compared each within-location infection rate from the Cladogenetic Migration SIR with the corresponding events in the resident-infects-resident class from the Visitor model. We also compared each between-location infection rate from the Cladogenetic Migration SIR model with the corresponding events in the visitor-infects-resident and resident-infects-visitor classes from the Visitor SIR model.

Finally, we used stochastic mapping for ancestral state estimation under both models in <monospace>RevBayes </monospace> (36). The stochastic mapping provides the sequence of transmission events for one realization of the tree under each model. We summarized the number of different transmission types by tabulating all events for each stochastic mapping generated by each Bayesian MCMC sample.

## Data and Code Availability

RevBayes analysis scripts, simulation scripts, plotting scrips, and simulated and empirical datasets are hosted through the public GitHub repository https://github.com/alberts2/Visitor-SSE.git.

## Supporting Information (SI)

We provide supplementary results and technical details of derivations in the main text in SI Appendix.

## ACKNOWLEDGMENTS

MJL is funded by the National Science Foundation (NSF Award DEB-2040347), the Fogarty International Center at the National Institutes of Health (Award Number R01 TW012704) as part of the joint NIH-NSF-NIFA Ecology and Evolution of Infectious Disease program, and the Washington University Incubator for Transdisciplinary Research. This research was supported in part by an appointment to the Department of Defense (DOD) Research Participation Program administered by the Oak Ridge Institute for Science and Education (ORISE) through an interagency agreement between the U.S. Department of Energy (DOE) and the DOD. ORISE is managed by ORAU under DOE contract number DE-SC0014664. All opinions expressed in this paper are the author’s and do not necessarily reflect the policies and views of DOD, DOE, or ORAU/ORISE. We are grateful to members of the Landis lab at Washington University in Saint Louis for constructive feedback on the manuscript.

