## Supplementary material for "Inferring epidemiological parameters under an infectious phylogeography model with visitor dynamics"

### Supplementary Information for: Inferring epidemiological parameters under an infectious phylogeography model with visitor dynamics

#### Contents

**S1 Table of notations** **S2**

**S2 Visitor SIR model** **S3**

S2.1 Approximating Visitor SIR model . . . . . S3

S2.2 Visitor SIR model state space . . . . . S6

S2.3 Anagenetic rates . . . . . S6

S2.4 Cladogenetic rates . . . . . S6

S2.5 Accuracy of the approximating model . . . . . S7

**S3 Cladogenetic Migration SIR model** **S9**

S3.1 Cladogenetic Migration SIR state space . . . . . S9

S3.2 Anagenetic rates . . . . . S10

S3.3 Cladogenetic rates . . . . . S10

**S4 Simple Migration SIR model** **S10**

**S5 Model comparison** **S11**

S5.1 Defining within-location and between-location infection rates . . . . . S11

S5.2 Simulated dataset using Visitor SIR model with three locations . . . . . S11

S5.3 Simulated dataset using Visitor SIR model with four locations . . . . . S16

S5.4 Simulated dataset using Visitor SIR model with five locations . . . . . S21

**S6 Transforming visitor state space for empirical analysis** **S27**

**S7 Countries with missing trips and nights data** **S28**

**S8 Ancestral state reconstruction under the Visitor SIR model** **S29**

S8.1 Labeled by home locations . . . . . S29

S8.2 Labeled by current locations . . . . . S29

S8.3 Labeled by matched locations . . . . . S30

### S1 Table of notations

**Table S1: A comprehensive list of notations used on this manuscript and their description.**

| Symbol | Description |
|---|---|
| $\mathcal{N}$ | Total population size in SIR structure |
| $S$ | Total susceptible population in SIR structure |
| $I$ | Total infected population in SIR structure |
| $R$ | Total recovered population in SIR structure |
| $\mathcal{N}_j$ | Total sub-population size in location $j$ |
| $N_j$ | Population size of location $j$ |
| $S_{ij}$ | Total number of susceptible individuals with home location $i$ and currently in location $j$ |
| $I_{ij}$ | Total number of infected individuals with home location $i$ and currently in location $j$ |
| $R_0$ | Basic reproduction number |
| $R_{0_i}$ | Basic reproduction number in location $i$ |
| $\delta_i$ | Per-capita sampling rate in location $i$ |
| $s_i$ | Sampling proportion in location $i$ |
| $\beta$ | Per-capita infection rate |
| $\beta_j$ | Per-capita infection rate in location $j$ |
| $\gamma$ | Per-capita recovery rate |
| $v_{i\to j}$ | Per-capita visit depart rate from home location $i$ to away location $j$ |
| $v^e_{i\to j}$ | Empirical per-capita visit depart rate from home location $i$ to away location $j$ |
| $r_{j\to i}$ | Per-capita visit return rate from current location $j$ to home location $i$ |
| $r^e_{j\to i}$ | Empirical per-capita visit return rate from current location $j$ to home location $i$ |
| $m_{i\to j}$ | Per-capita migration rate from location $i$ to location $j$ |
| $\mathbf{V}_i$ | Visitor matrix for an individual with home location $i$ under the Visitor SIR model |
| $\boldsymbol{P}_i$ | Stationary distribution of susceptible population in location $i$ |
| $\beta'_{hj}$ | Adjusted per-capita infection rate in location $j$ for a susceptible individual<br>with home location $h$ |
| $\mathcal{H}$ | Set of discrete home locations in the Visitor SIR model |
| $\mathcal{A}$ | Set of discrete away locations in the Visitor SIR model |
| $N$ | Total number of locations |
| $\mathcal{S}$ | Compound state space in the Visitor SIR model |
| $S_e$ | Empirical state space from Nadeau et al. (2021) |
| $\hat{S}_e$ | Transformed state space from Nadeau et al. (2021) with ambiguous locations |
| $\lambda_{(h,j),(i,j)}$ | Per-capita infection rate in location $j$ for a susceptible individual with home location $i$<br>by an infectious individual from home location $h$ under the approximation model of Visitor SIR |
| $\mathcal{C}$ | Cladogenetic rate matrix for the Visitor SIR model |
| $\mathcal{M}$ | Anagenetic rate matrix for the Visitor SIR model |
| $\lambda^0_{ij}$ | Per-capita infection rate in location $i$ under Cladogenetic Migration SIR<br>and Simple Migration SIR models |
| $p$ | Cladogenetic probability of observing no change in location following an infection event |
| $\hat{\lambda}_{ij}$ | Per-capita infection rate in location $j$ from an infected individual migrating<br>from location $i$ under Cladogenetic Migration SIR and Simple Migration SIR models |

| | |
|---|---|
| $\mathcal{L}$ | Set of of discrete locations in both Cladogenetic Migration SIR and Simple Migration SIR models |
| $\mathbb{N}$ | Set of all natural numbers |
| $\hat{\mathcal{M}}$ | Anagenetic rate matrix for the Cladogenetic Migration SIR and Simple Migration SIR models |
| $\hat{\mathcal{C}}$ | Cladogenetic rate matrix for the Cladogenetic Migration SIR and Simple Migration SIR models |
| Lognormal$(\mu, \sigma)$ | Log normal distribution with mean $\mu$ and standard deviation $\sigma$ |
| U$(a, b)$ | Uniform distribution with lower bound $a$ and upper bound $b$ |
| LogU$(a, b)$ | Log uniform distribution with lower bound $a$ and upper bound $b$ |
| $E[t_w]$ | Expected waiting time until a within-location infection occurs |
| $\hat{E}[t_b]$ | Expected waiting time until a between-location infection occurs in Cladogenetic Migration SIR model |
| $E[t_b]$ | Expected waiting time until a between-location infection occurs in Visitor SIR model |

### S2 Visitor SIR model

We define transmission dynamics under the Visitor SIR model. Suppose we have a constant population size $\mathcal{N}$ that consists of a susceptible compartment $S$, and infected compartment $I$, and recovered compartment $R$. We assume that an infected individual transitions to the recovered compartment upon sampling with some rate $\delta$. Under our model, $S$ and $I$ compartments can be sub-divided based on each individual's home location $i$ and current location $j$. That is we have,

$$S = \sum_i \sum_j S_{ij}, \tag{1}$$

$$I = \sum_i \sum_j I_{ij}. \tag{2}$$

Furthermore, we define a visitor as an individual (susceptible or infectious) from a home location $i$ that is currently visiting a different location $j$ where $i \neq j$. We define a resident as an individual from a home location $i$ that is currently in a location $j$ where $i = j$. There are four ways for an infectious individual to transmit a disease to a susceptible individual, namely resident-infects-resident, resident-infects-visitor, visitor-infects-resident, and visitor-infects-visitor types (See Eqs. (3)-(6)).

$$S_{jj} + I_{jj} \xrightarrow{\beta_j/\mathcal{N}_j} 2I_{jj} \qquad \text{A resident from } j \text{ infects another resident from } j \text{ in } j \tag{3}$$

$$S_{ij} + I_{jj} \xrightarrow{\beta_j/\mathcal{N}_j} I_{ij} + I_{jj} \qquad \text{A visitor from } i \text{ gets infected by a resident from } j \text{ in } j \tag{4}$$

$$S_{jj} + I_{ij} \xrightarrow{\beta_j/\mathcal{N}_j} I_{jj} + I_{ij} \qquad \text{A resident from } j \text{ gets infected by visitor from } i \text{ in } j \tag{5}$$

$$S_{ij} + I_{kj} \xrightarrow{\beta_j/\mathcal{N}_j} I_{ij} + I_{kj} \qquad \text{A visitor from } i \text{ gets infected by a visitor from } k \text{ in } j \tag{6}$$

where $\beta_j$ is the infection rate in location $j$, and $\mathcal{N}_j$ is the total number of individuals in location $j$.

##### S2.1 Approximating Visitor SIR model

Here we describe how to approximate the full process under the Visitor SIR model. Assuming the rates of visiting and returning are high relative to the rate at which the susceptible pool is draining, $\frac{\beta}{\mathcal{N}} \times S \times I$, then the distribution of the susceptible population, $S$, should be relatively close to its stationary

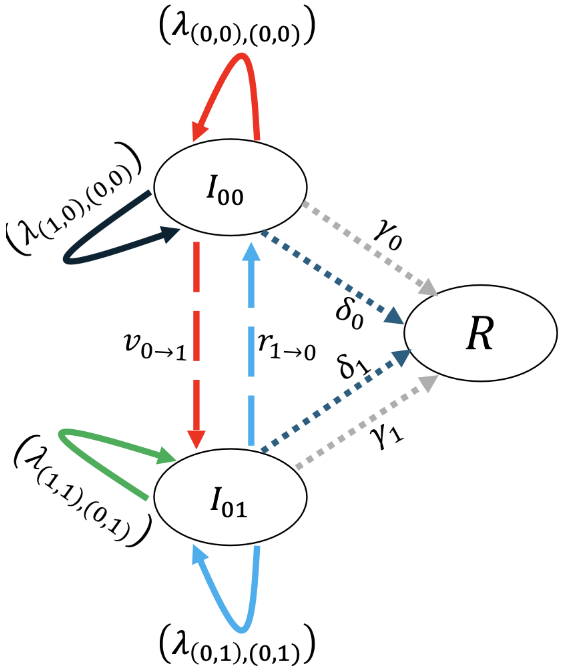


**Figure S1: Illustration of transmission dynamics under the approximated Visitor SIR model, viewed from the perspective of a susceptible individual with home location in 0.** By symmetry, the transmission dynamics for a susceptible individual with home location in 1 follows in the same manner. An arrow entering a compartment corresponds to gaining a new infection in that compartment. For example, the red arrow entering $I_{00}$ compartment means we have a newly infected individual with home location 0 and currently in the same location. The brackets represent the approximated per-capita infection rate where the first index indicates the infectious individual's home and current locations that causes a new infection, and the second index indicates the newly infected individual's hone and current locations. In this illustration, the solid red arrow corresponds to a resident-infects-resident infection event (See Fig. 1(b)), the solid **black** arrow corresponds to a visitor-infects-resident infection event (See Fig. 1(c)), the solid green arrow corresponds to a resident-infects-visitor infection event (See Fig. 1(d)), and the solid blue arrow corresponds to a visitor-infects-visitor infection event (See Fig. 1(e)). The dashed arrows corresponds to anagenetic events (host movement). The dotted arrows corresponds to both the recovery rates (dotted gray) and sampling rates (dotted blue) in each current location.

distribution. That is we have,

$$\frac{ds_{ij}(t)}{dt} = 0, \tag{7}$$

$$s_{ij} = \frac{S_{ij}(t)}{\sum_j S_{ij}(t)}. \tag{8}$$

Given this assumption, we no longer need to track the movement of susceptible individuals within their respective subpopulations (Fig. S1). To compute the stationary distribution of susceptible population size in each location, we construct a visitor matrix for each location. The visitor matrix for all locations is reducible to visitor matrices for each location if we disallow people visiting locations from other locations they are currently visiting. We denote these rates with $v_{i\to j}$ which is the per-capita visit depart rate for an individual to move from home location $i$ to away location $j$. Then, we denote the return rates to home location $i$ from any away location $j$ with $r_{j\to i}$. For example, the visitor matrix $\mathbf{V}_i$ for someone with home location $i$ is given by,

$$\mathbf{V}_i = \begin{bmatrix} -\sum_j v_{i\to j} & v_{i\to 2} & v_{i\to 3} & \cdots \\ r_{2\to i} & -r_{2\to i} & 0 & \\ r_{3\to i} & 0 & -r_{3\to i} & \\ \vdots & & & \ddots \end{bmatrix}. \tag{9}$$

We get the stationary distribution of susceptible population in location $i$ by finding the eigenvector with zero eigenvalue from the matrix above. The left eigenvector with eigenvalue $= 0$ is

$$\boldsymbol{P}_i = \begin{bmatrix} \frac{\prod_{k\neq i} r_{k\to i}}{R_i} \\ \frac{v_{i\to 2}\prod_{k\notin\{i,2\}} r_{k\to i}}{R_i} \\ \frac{v_{i\to 3}\prod_{k\notin\{i,3\}} r_{k\to i}}{R_i} \\ \vdots \end{bmatrix}. \tag{10}$$

In other words, the numerator for the home location ($j = i$; first element in equation 10) is the product of return rates from each away location $k$ to location $i$. And the numerator for each visited location ($j \neq i$) is the product of the visit rate for that location and the return rates of all the other locations. The denominator, $R_i$, is the sum of the numerators to make the vector for location $i$ sum to 1:

$$R_i = \prod_{k\neq i}^{N_i} r_{k\to i} + \sum_{j\neq i}^{N_i} \left[ v_{i\to j} \prod_{k\notin\{i,j\}}^{N_i} r_{k\to i} \right]$$

In general, across all locations $i$, we have $P_{ij} = [\boldsymbol{P}_i]_j$.

We use Eq. (10) to find an appropriate adjusted infection rate, $\beta'_{ij}$ for $\beta_j$. For a given location, $j$, being visited, we approximate the process with direct infections of susceptible individuals *from* location i, $S_i$, by infected individuals *in* location $j$, $I_{kj}$. Remember that $i$ is the home (from) index, $j$ is the current (visiting) index, and $k$ is for indexing over all locations of infectious people also visiting location $j$. That is, we approximate the following differential equation from the full model

$$\frac{dI_{ij}}{dt} = \tfrac{\beta_j}{N_j} S_{ij} \textstyle\sum_k I_{kj} - \gamma I_{ij} \tag{11}$$

with the following equation

$$\frac{dI'_{ij}}{dt} = \tfrac{\beta'_{ij}}{N_j} S_i \textstyle\sum_k I_{kj} - \gamma I_{ij}. \tag{12}$$

Then, by setting $\frac{dI_{ij}}{dt} = \frac{dI'_{ij}}{dt}$ we have

$$\frac{dI_{ij}}{dt} = \frac{dI'_{ij}}{dt}, \tag{13}$$

$$\beta'_{ij} S_i = \beta_j S_{ij}, \tag{14}$$

$$\beta'_{ij} = \beta_j \frac{S_{ij}}{S_i}, \tag{15}$$

where $S_i = \sum_j S_{ij}(t)$. If we assume the visitor CTMC is at stationarity, then using Eq. (10), we have $\frac{S_{ij}}{S_i} = \frac{v_{i\to j}\prod_{k\notin\{i,j\}} r_{k\to i}}{R_i}$ and

$$\beta'_{ij} = \begin{cases} \beta_i \times \frac{\prod_{k\neq i} r_{k\to i}}{R_i} & \text{if } i = j \\ \beta_j \times \frac{v_{i\to j}\prod_{k\notin\{i,j\}} r_{k\to i}}{R_i} & \text{if } i \neq j \end{cases}. \tag{16}$$

This approximation from Eq. (16) replaces the infection rate vector $\boldsymbol{\beta}$ of length $N$ with an $N \times N$ infection rate matrix. It also replaces the susceptible location matrix $\boldsymbol{S}$ of size $N \times N$ with a vector of length $N$. From here on, we also define the following,

$$\lambda_{(h,j),(i,j)} = \frac{\beta'_{ij}}{N_j} S_i \approx \frac{\beta_j}{N_j} S_{ij} \tag{17}$$

as the per-capita infection rate in location $j$ for a susceptible individual with home location $i$ by an infectious individual from home location $h$ under the approximation model of Visitor SIR.

##### S2.2 Visitor SIR model state space

Suppose $\mathcal{H}$ represents a set of home locations and $\mathcal{A}$ represents a set of away locations such that $N = |\mathcal{H}| + |\mathcal{A}|$ (that is, $\mathcal{H} \cap \mathcal{A} = \emptyset$), where $N$ is the total number of locations. The state of the model is defined to be the joint combination of an individual's home-away locations, $(i, j)$, where $i$ indicates the individual's home location and $j$ indicates the individual's current location. Therefore, the compound state space $\mathcal{S}$ is of size, $|\mathcal{S}| = |\mathcal{H}| \times |\mathcal{A}|$.

In the following sections, we define events on a transmission tree under the Visitor SIR model in two parts. First, we describe anagenetic rates of change via the $\mathcal{M}$ matrix. Then, we describe how cladogenetic rates of change are computed through the $\mathcal{C}$ matrix.

##### S2.3 Anagenetic rates

In this section, we describe the movement of individuals through the movement matrix $\mathcal{M}$ where $i$ represents the "from state" and $j$ represents the "to state". Moreover, the subscripts $h$ and $a$ represent the individual's home and current locations. That is, we have compound states $\tilde{i} = (i_h, i_a)$ and $\tilde{j} = (j_h, j_a)$. For example, $\mathcal{M}_{(i_h,i_a),(j_h,j_a)}$ represents the movement of an individual with previous location pair $(i_h, i_a)$ to a new location pair $(j_h, j_a)$.

In general, $\mathcal{M}$ is of size $|\mathcal{S}| \times |\mathcal{S}|$. Each row of the matrix corresponds to an individual's home-current location, and each column of the matrix corresponds to the individual's new home-current location. In general, we can describe the matrix entries as follows,

$$\mathcal{M}_{(i_h,i_a),(j_h,j_a)} = \begin{cases} -\sum_{j_a} v_{i_h \to j_a}, \text{ if } i_h = j_h,\ i_a = j_a, h = a \\ -r_{i_a \to j_h} \text{ if } i_h = j_h,\ i_a = j_a, h \neq a \\ r_{i_a \to j_h}, \text{ if } i_h = j_h,\ i_h = j_a, \\ v_{i_h \to j_a}, \text{ if } i_h = j_h,\ i_h = i_a,\ i_h \neq j_a, \\ 0, \text{ if otherwise.} \end{cases} \tag{18}$$

For example, suppose we have $|\mathcal{H}| = 3$ home locations and $|\mathcal{A}| = 3$ away locations. Then, the movement matrix is given by,

$$\mathcal{M} = \left( \begin{array}{c|ccccccccc} & (1,1) & (1,2) & (1,3) & (2,1) & (2,2) & (2,3) & (3,1) & (3,2) & (3,3) \\ \hline (1,1) & - & v_{1\to 2} & v_{1\to 3} & 0 & 0 & 0 & 0 & 0 & 0 \\ (1,2) & r_{2\to 1} & - & 0 & 0 & 0 & 0 & 0 & 0 & 0 \\ (1,3) & r_{3\to 1} & 0 & - & 0 & 0 & 0 & 0 & 0 & 0 \\ (2,1) & 0 & 0 & 0 & - & r_{1\to 2} & 0 & 0 & 0 & 0 \\ (2,2) & 0 & 0 & 0 & v_{2\to 1} & - & v_{2\to 3} & 0 & 0 & 0 \\ (2,3) & 0 & 0 & 0 & 0 & r_{3\to 2} & - & 0 & 0 & 0 \\ (3,1) & 0 & 0 & 0 & 0 & 0 & 0 & - & 0 & r_{1\to 3} \\ (3,2) & 0 & 0 & 0 & 0 & 0 & 0 & 0 & - & r_{2\to 3} \\ (3,3) & 0 & 0 & 0 & 0 & 0 & 0 & v_{3\to 1} & v_{3\to 3} & - \end{array} \right).$$

##### S2.4 Cladogenetic rates

We also construct a cladogenetic rate matrix, $\mathcal{C}$. Each cladogenetic event inside this matrix has the following structure: $((ij),(ij),(kj))$ where the first $(ij)$ represents the state of the ancestral lineage, and the other $(ij)$ and $(kj)$ represent the two infected lineages (individuals) following a transmission event. $i$, $j$, and $k$ identify different locations in the model. Under our model, one of the new lineages always inherits the ancestral state, and a transmission event can occur due to one of the four possible cases: (1) resident-infects-resident where a infector in location $j$ with home in $j$ infects a susceptible resident who is also at home in location $j$ $(i = j = k)$, (2) visitor-infects-resident where an infector from home location $i$ who is currently visiting location $j$ infects a susceptible who is a resident in location $j$ $(k = j, i \neq k)$, (3) resident-infects-visitor where an infectious resident with home in location $j$ and is currently in $j$ infects a susceptible visitor from home location $k$ who is currently visiting location $j$ $(i = j, i \neq k)$, and (4) visitor-infects-visitor where an infector from home location $i$ who is currently visiting location $j$ infects a susceptible from home location $k$ who is also currently in location $j$ $(i \neq j, k \neq j)$.

For example, suppose we have $|\mathcal{H}| = 3$ home locations and $|\mathcal{A}| = 3$ away locations then we can define the cladogenetic rate matrix as follows,

$$\mathcal{C} = \begin{bmatrix} C^{11} & C^{12} & C^{13} \\ C^{21} & C^{22} & C^{23} \\ C^{31} & C^{32} & C^{33} \end{bmatrix}.$$

We can then describe the cladogenetic rates for an infected individual with state 11 (a resident with home and current locations 1) as a $9 \times 9$ matrix where the non-zero entries are as follows,

$$C^{11} = \left( \begin{array}{ll} (11,11,11) & = \lambda_{(1,1),(1,1)}, \text{ infecting a resident from 1} \\ (11,11,21) & = \frac{\lambda_{(1,1),(2,1)}}{2}, \text{ infecting a visitor from 2} \\ (11,21,11) & = \frac{\lambda_{(1,1),(2,1)}}{2}, \text{ infecting a visitor from 2} \\ (11,31,11) & = \frac{\lambda_{(1,1),(3,1)}}{2}, \text{ infecting a visitor from 3} \\ (11,11,31) & = \frac{\lambda_{(1,1),(3,1)}}{2}, \text{ infecting a visitor from 3} \end{array} \right),$$

where $\lambda_{(1,1),(1,1)}$, $\lambda_{(1,1),(2,1)}$ and $\lambda_{(1,1),(3,1)}$ are defined in Eq. (17). Likewise for $C^{22}$ and $C^{33}$.

Similarly, for an infected individual with state 12 (a visitor from home location 1 and currently in location 2), we can describe the cladogenetic event rates as a $9 \times 9$ matrix, $C^{12}$, where the non-zero entries are as follows,

$$C^{12} = \left( \begin{array}{ll} (12,12,22) & = \frac{\lambda_{(1,2),(2,2)}}{2}, \text{ infecting a resident from 2} \\ (12,22,12) & = \frac{\lambda_{(1,2),(2,2)}}{2}, \text{ infecting a resident from 2} \\ (12,12,12) & = \lambda_{(1,2),(1,2)}, \text{ infecting a visitor from 1} \\ (12,12,32) & = \frac{\lambda_{(1,2),(3,2)}}{2}, \text{ infecting a visitor from 3} \\ (12,32,12) & = \frac{\lambda_{(1,2),(3,2)}}{2}, \text{ infecting a visitor from 3} \end{array} \right),$$

where $\lambda_{(1,2),(1,2)}$, $\lambda_{(1,2),(2,2)}$ and $\lambda_{(1,2),(3,2)}$ are defined in Eq. (17). Likewise for $C^{13}$ and all the other off-diagonal block matrices in $C$. Moreover, we refer to a branching event without state change as a within-location infection under the visitor model e.g., $(11,11,11)$, and branching event with state change as a between-location infection e.g., $(11,11,21)$ that corresponds to resident-infects-visitor event and $(12,12,22)$ that corresponds to visitor-infects-resident event.

##### S2.5 Accuracy of the approximating model

We used MASTER's numerical method for approximating compartment size time series under the deterministic full and approximate visitor models. We found that the dynamics for the number of infectious individuals across different subpopulations match in both models (Figs. S2-S4).

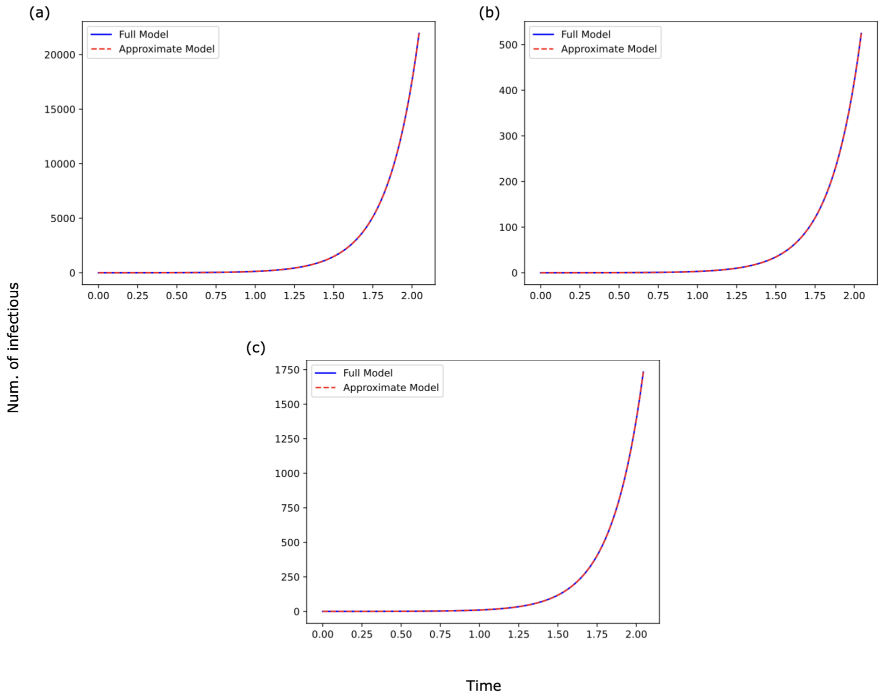


**Figure S2: Number of infections trajectory under the full model (solid blue) and the approximation (dashed red) with 3 locations $(\mathbf{0, 1, 2})$.** (a) compares the number of infections from home and away location 0, (b) compares for home location 1 and current location 0, and (c) compares for home location 2 and current location 0.

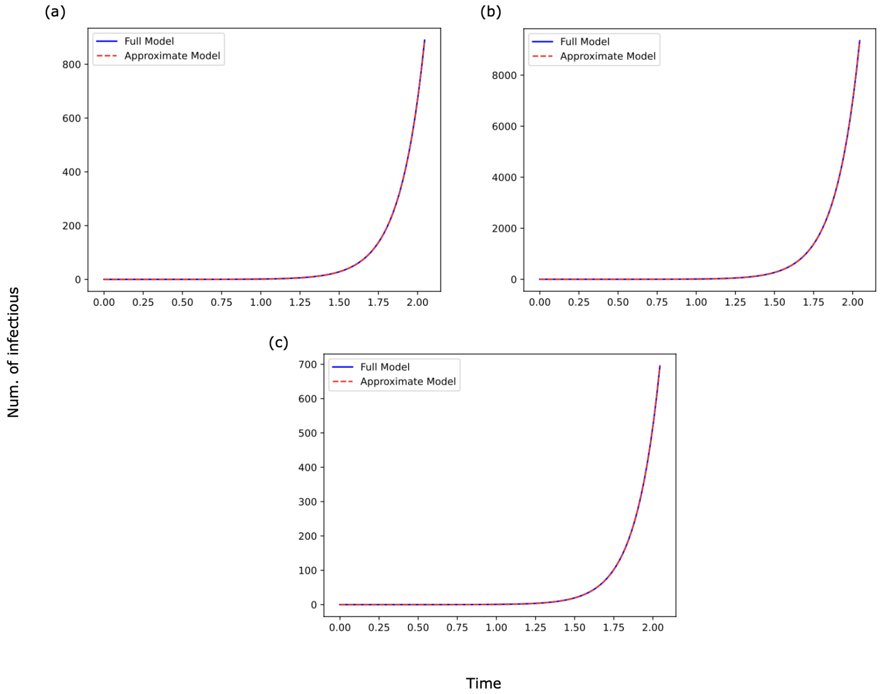


**Figure S3: Number of infections trajectory under the full model (solid blue) and the approximation (dashed red) with 3 locations $(\mathbf{0, 1, 2})$.** (a) compares the number of infections from home location 0 and current locations 1, (b) compares for home and away location 1, and (c) compares for home location 2 and current location 1.

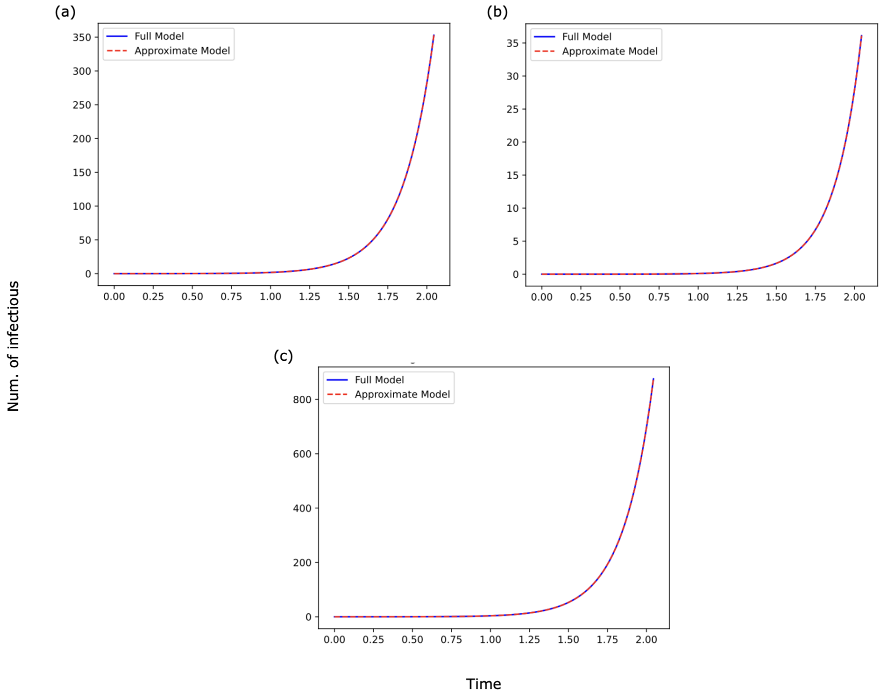


**Figure S4: Number of infections trajectory under the full model (solid blue) and the approximation (dashed red) with 3 locations $(0, 1, 2)$.** (a) compares the number of infections from home location 0 and current locations 2, (b) compares for home location 1 and current location 2, and (c) compares for home and away location 2.

### S3 Cladogenetic Migration SIR model

In this section, we define the standard SIR model with anagenesis (along branches) and cladogenesis (during branching). From here on, we refer to this model as the Cladogenetic Migration SIR model. In this model, the movement of each individual only depends on their current location. Assuming that we have equal rates among all $N$ locations, we define the per capita infection rate in location $i$ as follows.

$$\lambda^0_{ij} = (\delta_j + \gamma_j) \times R_{0_i}, \tag{19}$$

$$\hat{\lambda}_{ij} = \begin{cases} \lambda^0_{ii} \times p & \text{if } i = j \\ \lambda^0_{ij} \times (1-p) \times \frac{1}{N-1} & \text{if } i \neq j \end{cases}, \tag{20}$$

$$0 \leq p \leq 1, \tag{21}$$

where $\delta_i$ is the sampling rate in location $i$, $\gamma_i$ is the recovery rate in $i$, $R_{0_i}$ is the basic reproduction number in $i$, and $p$ is the cladogenetic probability of observing no change in location following an infection event, and $\frac{1}{N-1}$ is to take into account all possible locations where an infection could occur in the case of state (location) change.

#### S3.1 Cladogenetic Migration SIR state space

Suppose $\mathcal{L}$ represents a set of discrete locations (i.e., $|\mathcal{L}| \in \mathbb{N}$). The state of the model is the current location $i \in \mathcal{L}$ of an individual. Next, we define events on a transmission tree under the Cladogenetic Migration SIR model. First, we describe anagenetic rates of change via the $\hat{\mathcal{M}}$ matrix where its off-diagonal entries represent a host movement from the previous location to the current location. Then we describe how the cladogenetic rates of change are computed through the $\hat{\mathcal{C}}$ matrix. Each entry in this $\hat{\mathcal{C}}$

corresponds to an infection event in location $j$ of an infected individual as the individual moves from location $i$ to location $j$.

##### S3.2 Anagenetic rates

We describe the movement of an individual from location $i$ to a new location $j$ through the movement matrix $\hat{\mathcal{M}}$ where its size is equal to $|\mathcal{L}| \times |\mathcal{L}|$. Each row of the matrix corresponds to an individual's previous location, and each column corresponds to the current location. In general, we can describe the entries as follows.

$$\hat{\mathcal{M}}_{i,j} = \begin{cases} 0, & \text{if } i = j, \\ m_{i \to j}, & \text{if } i \neq j, \end{cases} \tag{22}$$

where $m_{i \to j}$ is migration rate from location $i$ to location $j$.

##### S3.3 Cladogenetic rates

We construct the cladogenetic rate matrix $\hat{\mathcal{C}}$ for the Cladogenetic Migration model. Each cladogenetic event is represented as $(i, j, k)$ where $i$ represents the previous location of the original infected individual, and $j$ represents the location where the new infection occurs. Similar to the visitor model, one of the child lineage always inherits the parent state, namely $i = k$. However, unlike the visitor model that tracks an individual's home and current locations, an infection event under the cladogenetic migration model occurs simultaneously with the movement of the infected individual.

For example, suppose we have $|\mathcal{L}| = 3$ locations then we can define the cladogenetic rate matrix as follows,

$$\hat{\mathcal{C}} = \begin{bmatrix} \hat{C}^1 & \hat{C}^2 & \hat{C}^3 \end{bmatrix}$$

We can then describe the cladogenetic rates for an infected individual with state $i = 1$ as a $3 \times 3$ matrix where the non-zero entries are as follows,

$$\hat{C}^1 = \begin{pmatrix} (1,1,1) & = \hat{\lambda}_{11} \\ (1,2,1) & = \frac{\hat{\lambda}_{12}}{2} \\ (1,1,2) & = \frac{\hat{\lambda}_{12}}{2} \\ (1,3,1) & = \frac{\hat{\lambda}_{13}}{2} \\ (1,1,3) & = \frac{\hat{\lambda}_{13}}{2} \end{pmatrix},$$

where $\hat{\lambda}_{ij}$ is defined in Eq. (20). Likewise, for $\hat{C}^2$ and $\hat{C}^3$. Moreover, we refer to a branching event without state change as a within-location infection e.g., $(1, 1, 1)$, and branching event with state change as a between-location infection e.g., $(1, 2, 1)$.

#### S4 Simple Migration SIR model

This is identical to the cladogenetic migration model except we do not allow cladogenesis during an infection event. In this model we simply have $\hat{\lambda}_{ij} = 0$ whenever $i \neq j$.

### S5 Model comparison

#### S5.1 Defining within-location and between-location infection rates

Besides comparing both $R_0$ and $\delta$ parameters across different models, we also compare the within-location and between-location infection rates. Since the Visitor SIR model has a different state space compared to the other two migration models, it is necessary to give a precise definition for both events. Under the Visitor SIR model, the within-location infection event is an infection to a susceptible resident by another infectious resident (See Fig. S5(b)). Under the Cladogenetic Migration SIR and Simple Migration SIR models, it is an infection event that occurs in the current location and does not involve movement of the newly infected individual (See Fig. S5(a)). Next, the between-location infection under the Visitor SIR model corresponds to resident-infects-visitor infection type where the newly infected individual returns to their home location (See Fig. S5(d)) or visitor-infects-resident infection type (See Fig. S5)(e) . For the Cladogenetic Migration SIR model, it is an infection in the current location that is immediately followed by a migration event of the newly infected individual to a different location (See Fig. S5(c)).

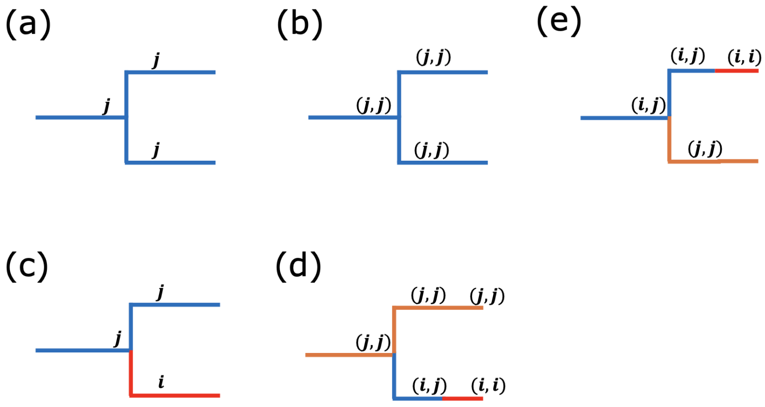


**Figure S5: Transmission modes under Visitor SIR and migration SIR models.** (a)-(b) within-location infection event under both migration models and Visitor SIR models, respectively; (c)-(e) between-location infection event under Cladogenetic Migration SIR and Visitor SIR, respectively.

#### S5.2 Simulated dataset using Visitor SIR model with three locations

Here, for each different relationship of visit depart and visit return as described in Table S2, we simulated 116 phylogenetic trees from the visitor SIR model with 3 locations. Then, we conducted a Bayesian coverage experiment using the same simulating model under Visitor SIR. Figure S6 shows the coverages levels across different parameters when fitted the the Visitor SIR model with three locations for 116 tranmission trees simulated under the same model. In summary, most of the parameters across different movement scenarios, as described in Table S2, have coverage within the expectation. The coverage $\log\left(\frac{v_{i\to j}}{r_{j\to i}}\right)$ in Figure S6(a) is lower than expected. We are not sure what caused this behavior. However, we see an improvement in the coverage for the same parameter with more locations (See Figs. S10−S14(a)). Next, comparing with the migration models, as seen in Figures S7−S9, the coverage for the within-location and between-location infection rates are always low in both migration models (Cladogenetic Migration SIR and Simple Migration SIR).

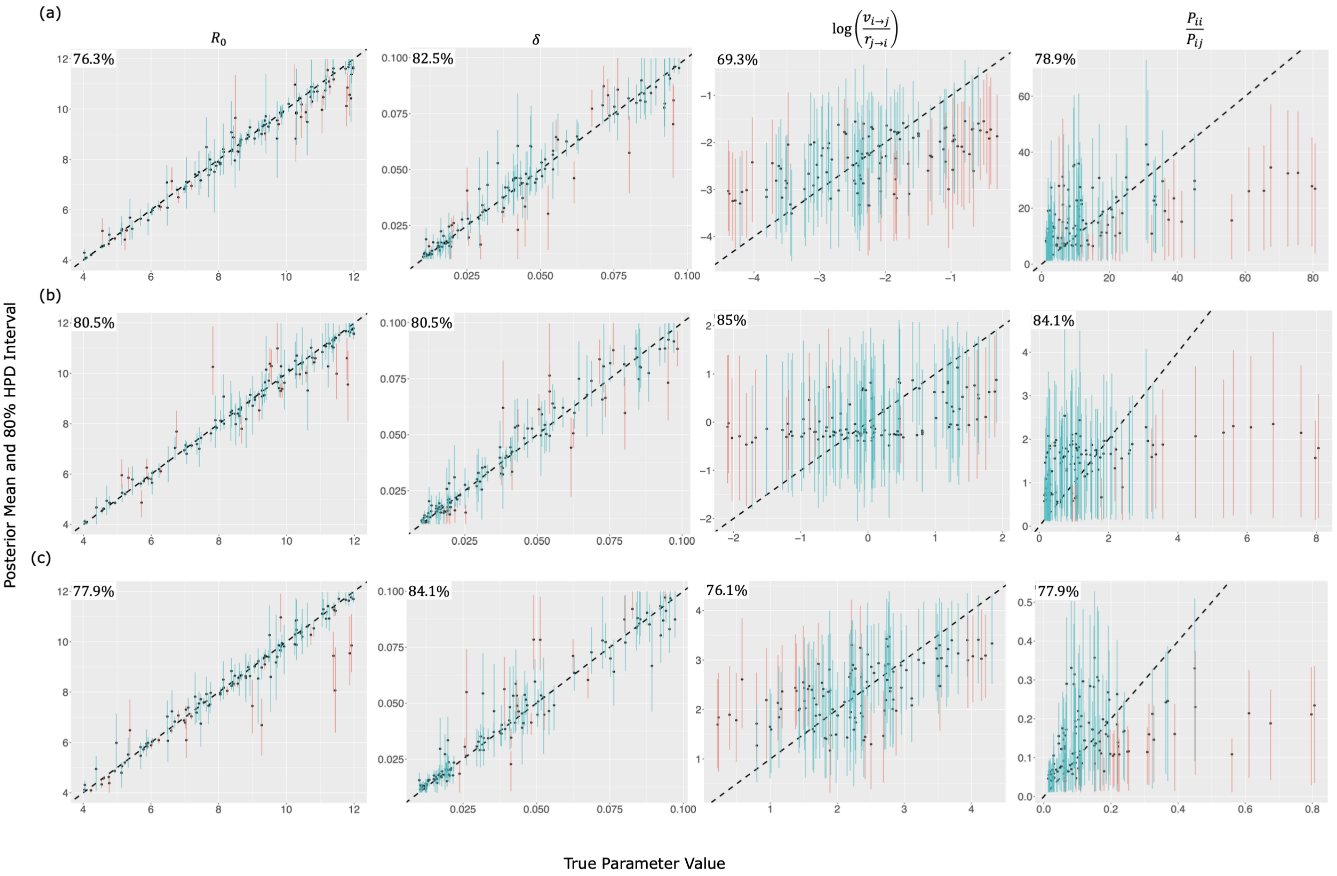


**Figure S6: Quality of parameter estimation and Bayesian coverage of the Visitor SIR model parameters in simulations with three locations and under all scenarios described in Table S2, namely (a) depart rate < return rate (top panel), (b) depart rate = return rate (middle panel), (c) depart rate > return rate (bottom panel).** Plots show true parameter values on the $x-$axis and estimated values on the $y-$axis. Parameter names are shown in the top of the upper plots, and the 80% HPD intervals, which cover the true values, are shown in blue. Intervals which do not cover the true values, are shown in red. The coverage percentages are shown in the top left of each plot. From these plots, we observe that some parameters can be inferred reasonably well.

**Table S2: Table showing priors for each estimated parameters with their definitions for each different model across three simulation scenarios under the Visitor SIR model.**

| Notation | Visitor SIR | Clado Migration SIR | Simple Migration SIR | Description |
|---|---|---|---|---|
| | Priors | | | |
| **(a) 3 locations, depart rate < return rate** | | | | |
| $R_0$ | U(4,12) | U(4,12) | U(4,12) | basic reproduction number |
| $\gamma$ | U(0.1,1) | U(0.1,1) | U(0.1,1) | recovery rate |
| $\delta$ | LogU(0.01,0.1) | LogU(0.01,0.1) | LogU(0.01,0.1) | sampling rate |
| $v_{i \to j}, \forall i, j$ | LogU(0.01,0.1) | – | – | depart rate |
| $r_{j \to i}, \forall i, j$ | LogU(0.1,1.0) | – | – | return rate |
| $m_{i \to j}, \forall i, j$ | – | LogU(0.01,0.1) | LogU(0.01,0.1) | migration rate |
| $p$ | – | U(0,1) | U(0,1) | Cladogenetic probability of observing no change in state |
| **(b) 3 locations, depart rate = return rate** | | | | |
| $R_0$ | U(4,12) | U(4,12) | U(4,12) | basic reproduction number |
| $\gamma$ | U(0.1,1) | U(0.1,1) | U(0.1,1) | recovery rate |
| $\delta$ | LogU(0.01,0.1) | LogU(0.01,0.1) | LogU(0.01,0.1) | sampling rate |
| $v_{i \to j}, \forall i, j$ | LogU(0.1,1.0) | – | – | depart rate |
| $r_{j \to i}, \forall i, j$ | LogU(0.1,1.0) | – | – | return rate |
| $m_{i \to j}, \forall i, j$ | – | LogU(0.1,1.0) | LogU(0.1,1.0) | migration rate |
| $p$ | – | U(0,1) | U(0,1) | Cladogenetic probability of observing no change in state |
| **(c) 3 locations, depart rate > return rate** | | | | |
| $R_0$ | U(4,12) | U(4,12) | U(4,12) | basic reproduction number |
| $\gamma$ | U(0.1,1) | U(0.1,1) | U(0.1,1) | recovery rate |
| $\delta$ | LogU(0.01,0.1) | LogU(0.01,0.1) | LogU(0.01,0.1) | sampling rate |
| $v_{i \to j}, \forall i, j$ | LogU(0.1,1.0) | – | – | depart rate |
| $r_{j \to i}, \forall i, j$ | LogU(0.01,0.1) | – | – | return rate |
| $m_{i \to j}, \forall i, j$ | – | LogU(0.1,1.0) | LogU(0.1,1.0) | migration rate |
| $p$ | – | U(0,1) | U(0,1) | Cladogenetic probability of observing no change in state |

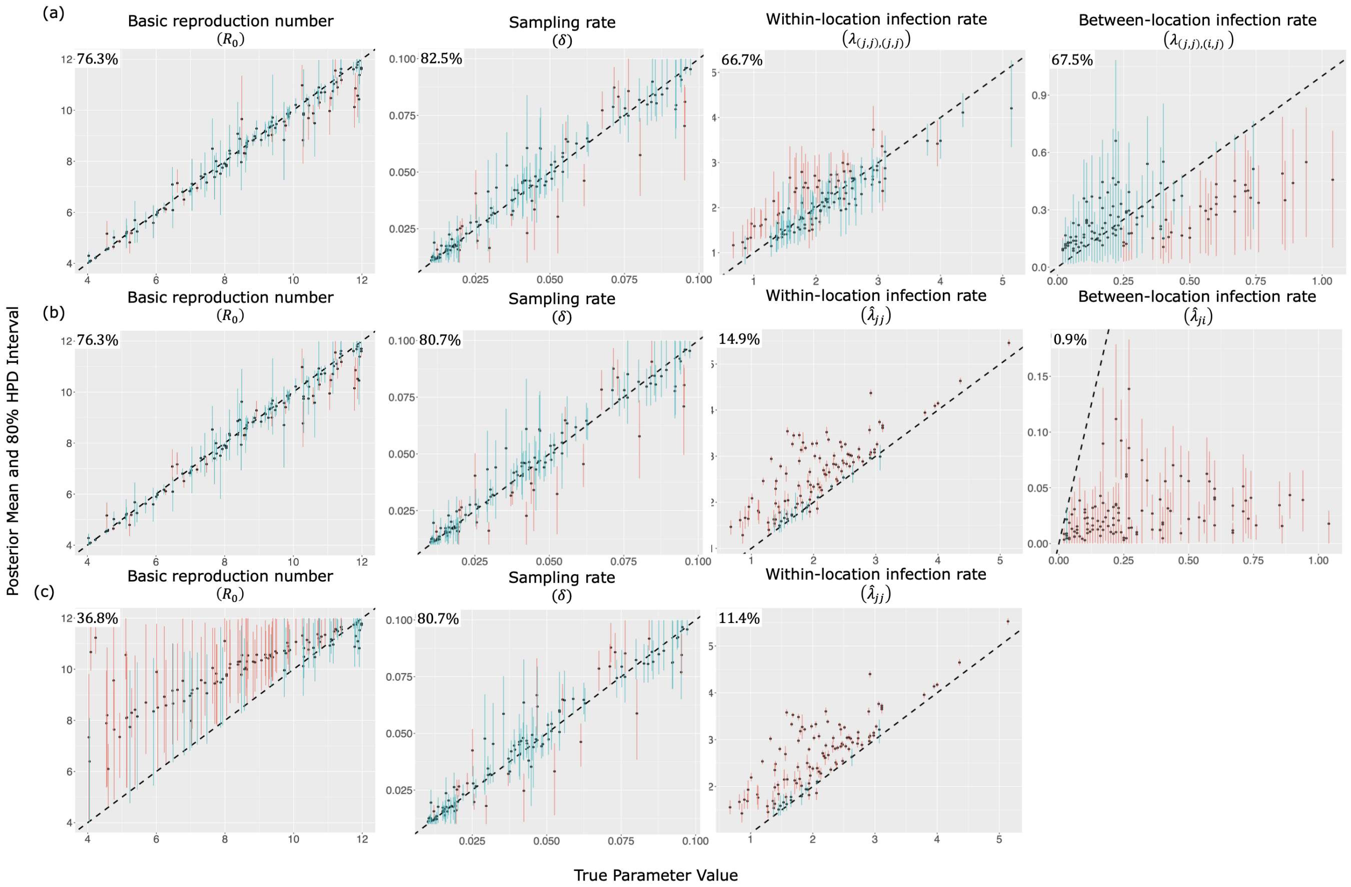


**Figure S7: Quality of parameter estimation and coverage across the inference models, namely (a) the Visitor SIR, (b) Cladogenetic Migration SIR, and (c) Simple Migration SIR models where the simulation model (the true model) is simulated under the same Visitor SIR model with 3 locations as the Visitor SIR model used for inference, as described in Table. S2(a).** Plots show true values on the $x-$axis and the estimated values on the $y-$axis. For each plot, 80% HPD intervals, which covers the truth, are shown in blue, and 80% intervals, which do not cover the truth, are shown in red. The coverage percentages are shown in the top left of each plot.

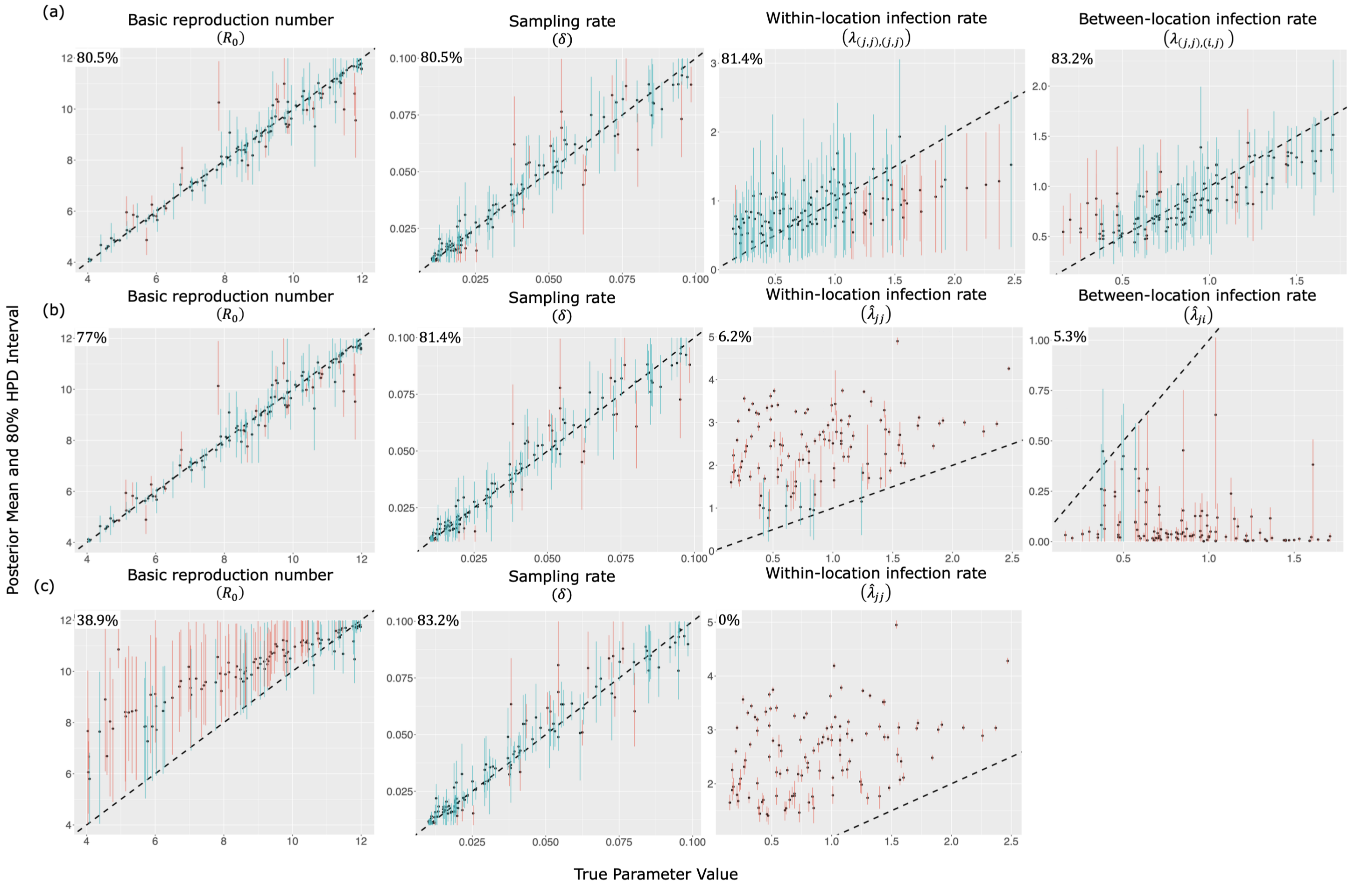


**Figure S8: Quality of parameter estimation and coverage across the inference models, namely (a) the Visitor SIR, (b) Cladogenetic Migration SIR, and (c) Simple Migration SIR models where the simulation model (the true model) is simulated under the same Visitor SIR model with 3 locations as the Visitor SIR model used for inference, as described in Table. S2(b).** Plots show true values on the $x-$axis and the estimated values on the $y-$axis. For each plot, 80% HPD intervals, which covers the truth, are shown in blue, and 80% intervals, which do not cover the truth, are shown in red. The coverage percentages are shown in the top left of each plot.

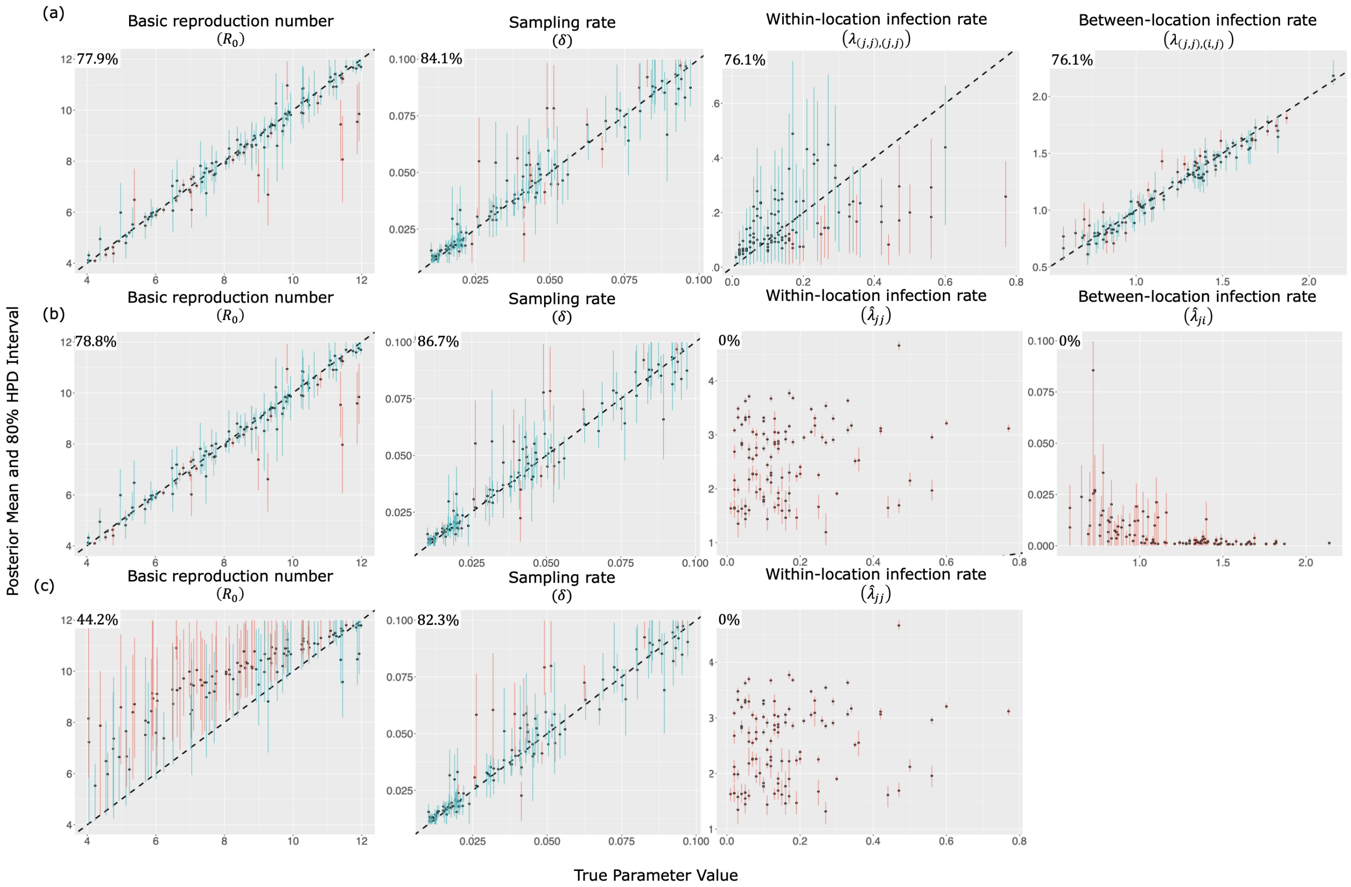


**Figure S9: Quality of parameter estimation and coverage across the inference models, namely (a) the Visitor SIR, (b) Cladogenetic Migration SIR, and (c) Simple Migration SIR models where the simulation model (the true model) is simulated under the same Visitor SIR model with 3 locations as the Visitor SIR model used for inference, as described in Table. S2(c).** Plots show true values on the $x-$axis and the estimated values on the $y-$axis. For each plot, 80% HPD intervals, which covers the truth, are shown in blue, and 80% intervals, which do not cover the truth, are shown in red. The coverage percentages are shown in the top left of each plot.

##### S5.3 Simulated dataset using Visitor SIR model with four locations

For each set of visit depart and visit return rates as described in Table S3, we simulated 116 phylogenetic trees from the Visitor SIR model with 4 locations. Then, we conducted a Bayesian coverage experiment using the same simulating model under Visitor SIR. Figure S10 shows the coverage levels across different parameters when fit to the the Visitor SIR model with four locations for the 116 trees simulated under the same model. In summary, most of the parameters across different movement scenarios, as described in Table S3, have coverages near expectation. One exception is the the coverage for $R_0$ in Figure S10(b) which seems lower. However, this may be caused by some simulated trees being past the exponential phase of disease transmission. Next, comparing with the migration models, as seen in Figures S11−S13, the coverage for the within-location and between-location infection rates are always low in both migration models (Cladogenetic Migration SIR and Simple Migration SIR).

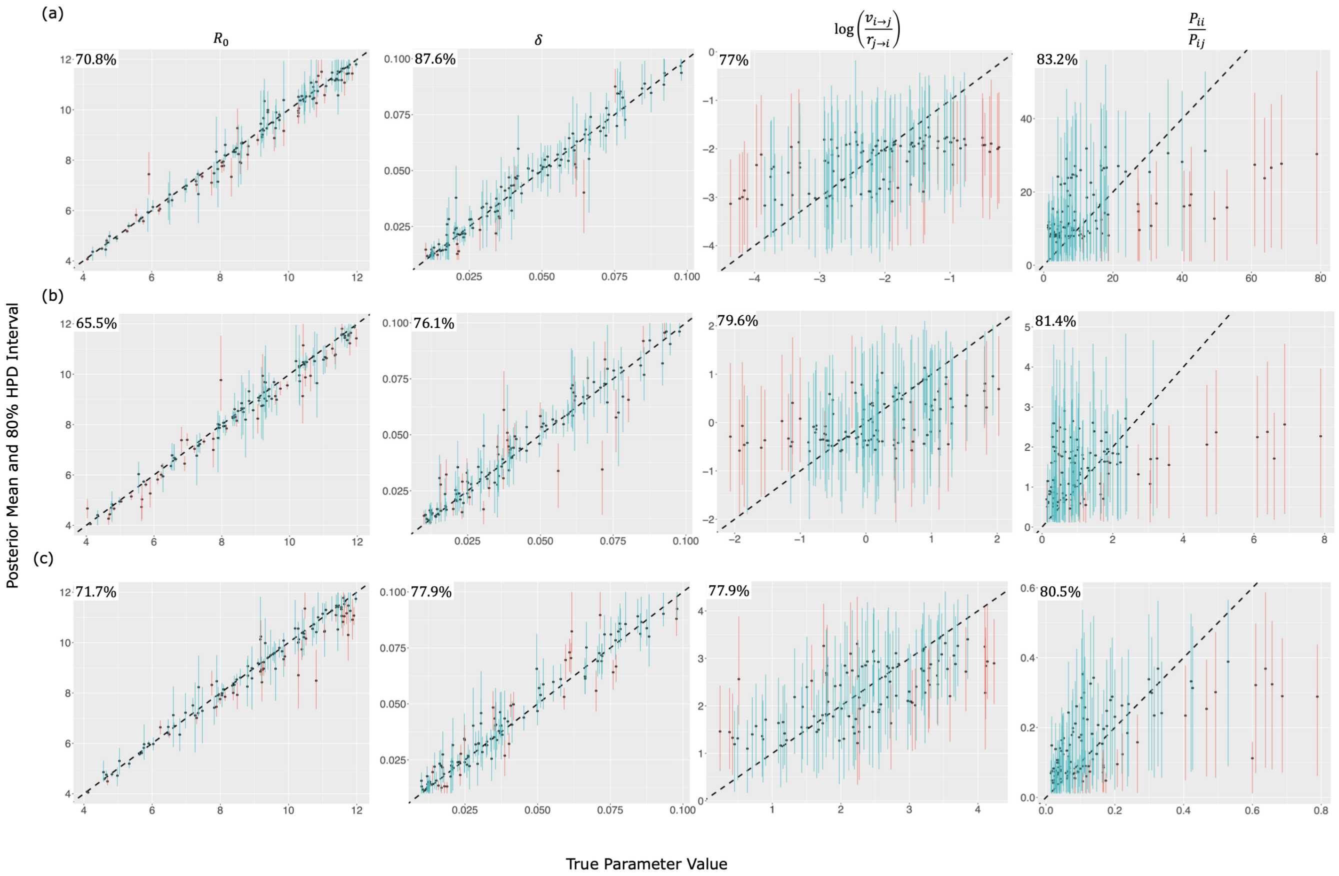


**Figure S10: Quality of parameter estimation and Bayesian coverage of the Visitor SIR model parameters in simulations with four locations and scenarios described in Table S3, namely (a) depart rate < return rate (top panel), (b) depart rate = return rate (middle panel), (c) depart rate > return rate (bottom panel).** Plots show true parameter values on the $x-$axis and estimated values on the $y-$axis. Parameter names are shown in the top of the upper plots, and the 80% HPD intervals, which cover the true values, are shown in blue, and the intervals, which do not cover the true values, are shown in red. The coverage percentages are shown in the top left of each plot.

**Table S3: Table showing priors for each estimated parameters with their definitions for each different model across three simulation scenarios under the Visitor SIR model.**

| Notation | Visitor SIR | Clado Migration SIR | Simple Migration SIR | Description |
|---|---|---|---|---|
| | Priors | | | |
| **(a) 4 locations, depart rate < return rate** | | | | |
| $R_0$ | U(4,12) | U(4,12) | U(4,12) | basic reproduction number |
| $\gamma$ | U(0.1,1) | U(0.1,1) | U(0.1,1) | recovery rate |
| $\delta$ | LogU(0.01,0.1) | LogU(0.01,0.1) | LogU(0.01,0.1) | sampling rate |
| $v_{i\to j}, \forall i,j$ | $\text{LogU}\left(\frac{2(0.01)}{3}, \frac{2(0.1)}{3}\right)$ | – | – | depart rate |
| $r_{j\to i}, \forall i,j$ | $\text{LogU}\left(\frac{2(0.1)}{3}, \frac{2(1.0)}{3}\right)$ | – | – | return rate |
| $m_{i\to j}, \forall i,j$ | – | $\text{LogU}\left(\frac{2(0.01)}{3}, \frac{2(0.1)}{3}\right)$ | $\text{LogU}\left(\frac{2(0.01)}{3}, \frac{2(0.1)}{3}\right)$ | migration rate |
| $p$ | – | U(0,1) | U(0,1) | Cladogenetic probability of observing no change in state |
| **(b) 4 locations, depart rate = return rate** | | | | |
| $R_0$ | U(4,12) | U(4,12) | U(4,12) | basic reproduction number |
| $\gamma$ | U(0.1,1) | U(0.1,1) | U(0.1,1) | recovery rate |
| $\delta$ | LogU(0.01,0.1) | LogU(0.01,0.1) | LogU(0.01,0.1) | sampling rate |
| $v_{i\to j}, \forall i,j$ | $\text{LogU}\left(\frac{2(0.1)}{3}, \frac{2(1.0)}{3}\right)$ | – | – | depart rate |
| $r_{j\to i}, \forall i,j$ | $\text{LogU}\left(\frac{2(0.1)}{3}, \frac{2(1.0)}{3}\right)$ | – | – | return rate |
| $m_{i\to j}, \forall i,j$ | – | $\text{LogU}\left(\frac{2(0.1)}{3}, \frac{2(1.0)}{3}\right)$ | $\text{LogU}\left(\frac{2(0.1)}{3}, \frac{2(1.0)}{3}\right)$ | migration rate |
| $p$ | – | U(0,1) | U(0,1) | Cladogenetic probability of observing no change in state |
| **(c) 4 locations, depart rate > return rate** | | | | |
| $R_0$ | U(4,12) | U(4,12) | U(4,12) | basic reproduction number |
| $\gamma$ | U(0.1,1) | U(0.1,1) | U(0.1,1) | recovery rate |
| $\delta$ | LogU(0.01,0.1) | LogU(0.01,0.1) | LogU(0.01,0.1) | sampling rate |
| $v_{i\to j}, \forall i,j$ | $\text{LogU}\left(\frac{2(0.1)}{3}, \frac{2(1.0)}{3}\right)$ | – | – | depart rate |
| $r_{j\to i}, \forall i,j$ | $\text{LogU}\left(\frac{2(0.01)}{3}, \frac{2(0.1)}{3}\right)$ | – | – | return rate |
| $m_{i\to j}, \forall i,j$ | – | $\text{LogU}\left(\frac{2(0.1)}{3}, \frac{2(1.0)}{3}\right)$ | $\text{LogU}\left(\frac{2(0.1)}{3}, \frac{2(1.0)}{3}\right)$ | migration rate |
| $p$ | – | U(0,1) | U(0,1) | Cladogenetic probability of observing no change in state |

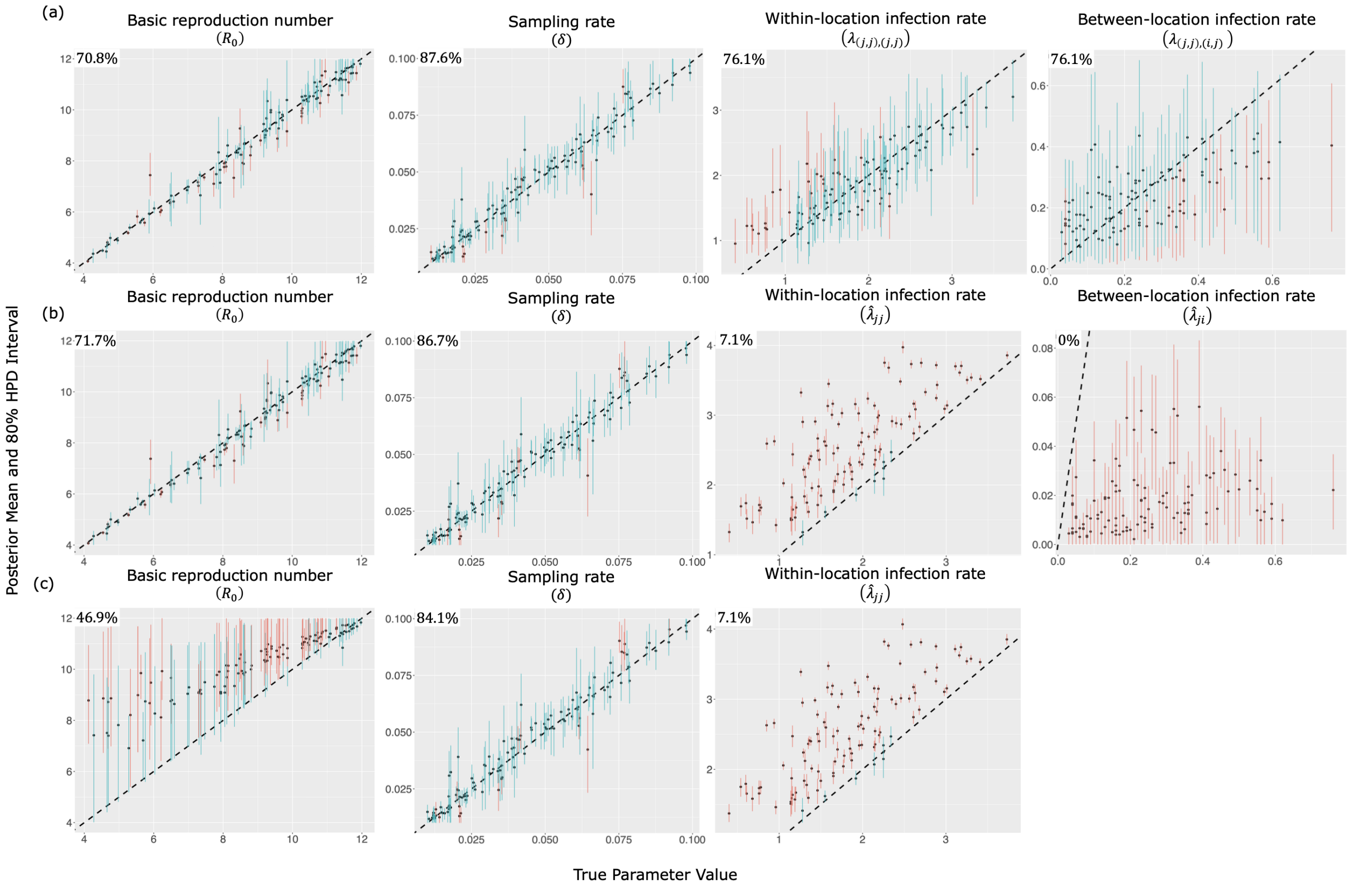


**Figure S11: Quality of parameter estimation and coverage across the inference models, namely (a) the Visitor SIR, (b) Cladogenetic Migration SIR, and (c) Simple Migration SIR models where the simulation model (the true model) is simulated under the same Visitor SIR model with 4 locations as the Visitor SIR model used for inference, as described in Table. S3(a).** Plots show true values on the $x-$axis and the estimated values on the $y-$axis. For each plot, 80% HPD intervals, which covers the truth, are shown in blue, and 80% intervals, which do not cover the truth, are shown in red. The coverage percentages are shown in the top left of each plot.

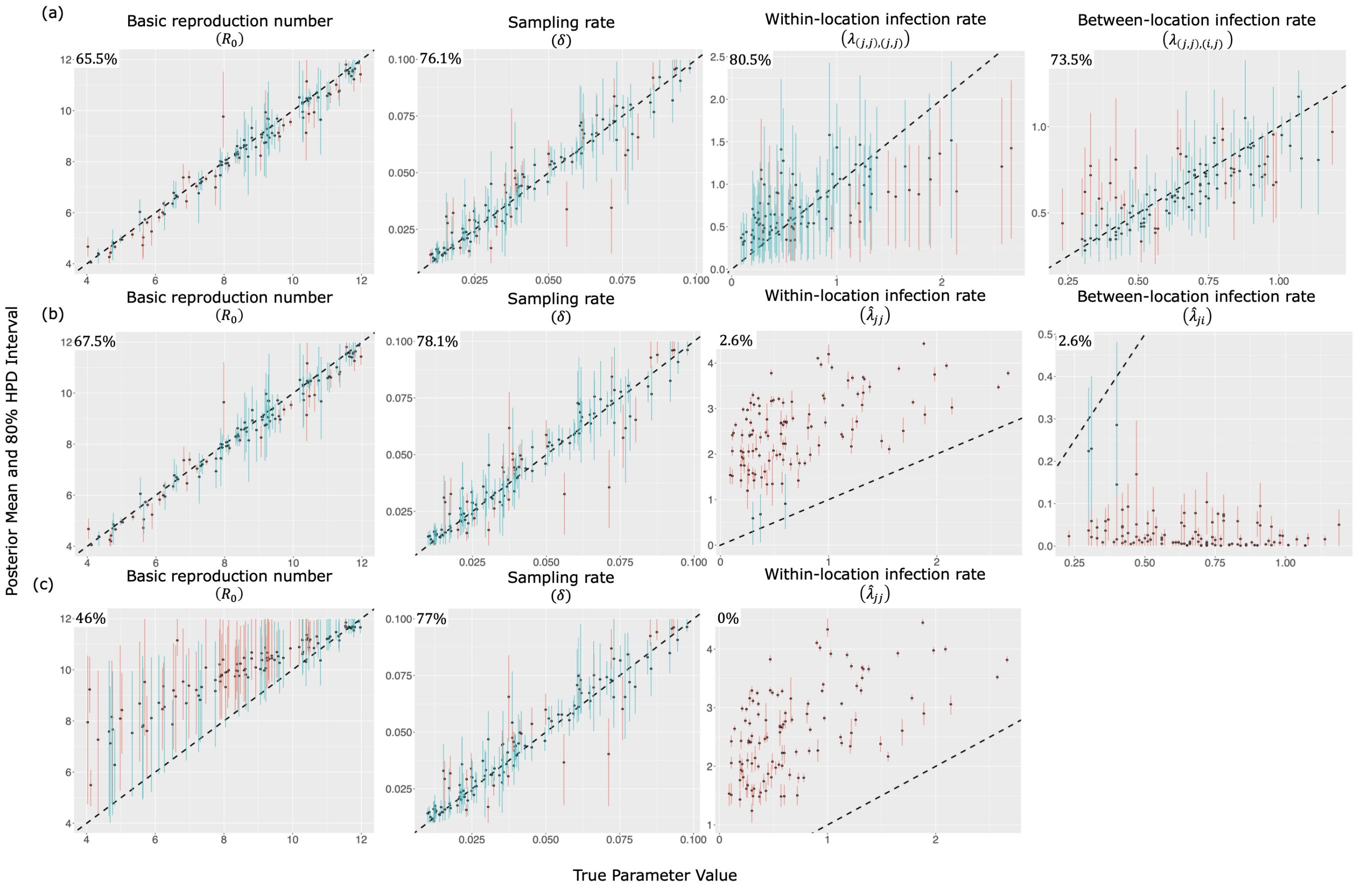


**Figure S12: Quality of parameter estimation and coverage across the inference models, namely (a) the Visitor SIR, (b) Cladogenetic Migration SIR, and (c) Simple Migration SIR models where the simulation model (the true model) is simulated under the same Visitor SIR model with 4 locations as the Visitor SIR model used for inference, as described in Table. S3(b).** Plots show true values on the $x-$axis and the estimated values on the $y-$axis. For each plot, 80% HPD intervals, which covers the truth, are shown in blue, and 80% intervals, which do not cover the truth, are shown in red. The coverage percentages are shown in the top left of each plot.

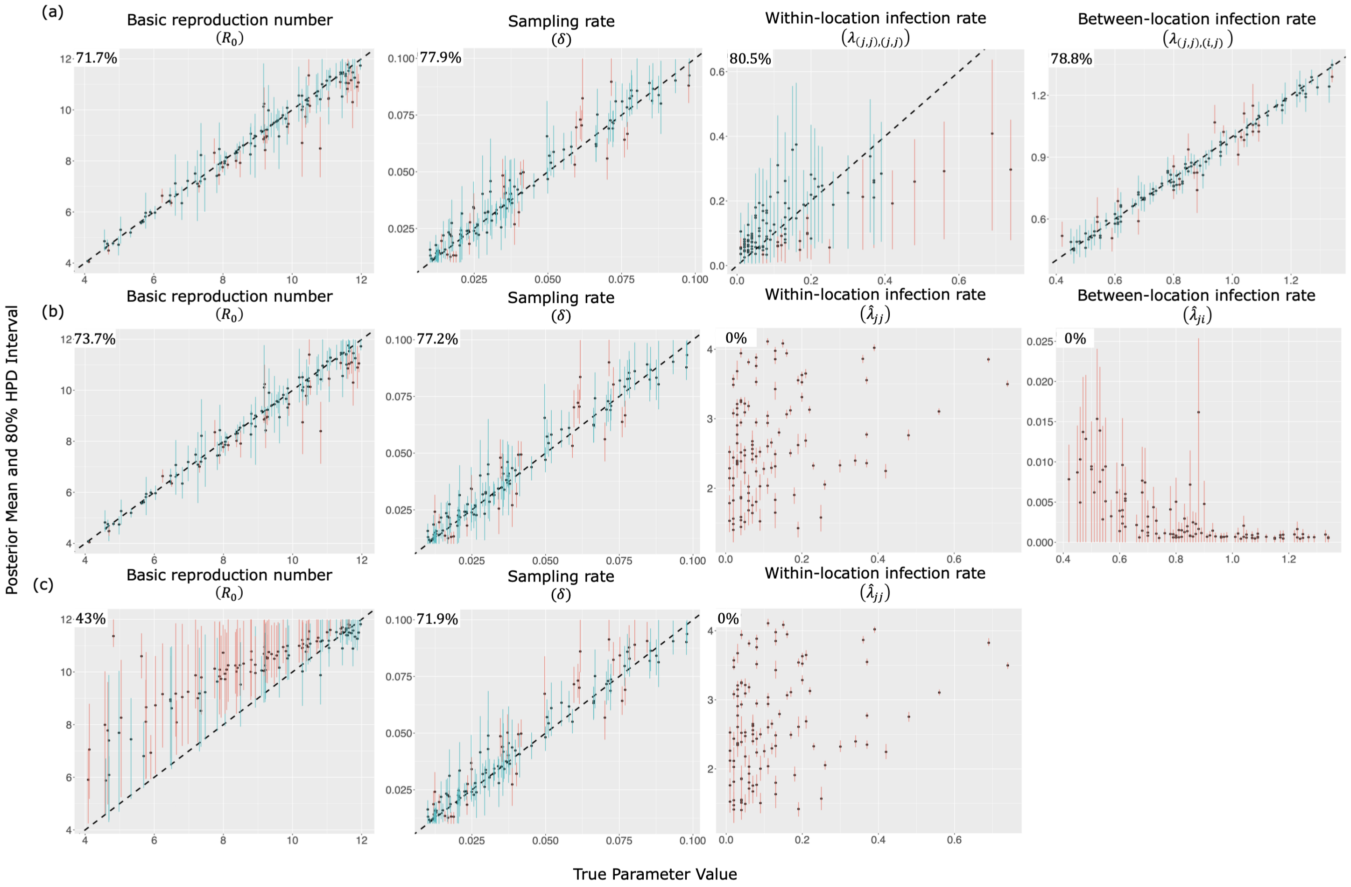


**Figure S13: Quality of parameter estimation and coverage across the inference models, namely (a) the Visitor SIR, (b) Cladogenetic Migration SIR, and (c) Simple Migration SIR models where the simulation model (the true model) is simulated under the same Visitor SIR model with 4 locations as the Visitor SIR model used for inference, as described in Table. S3(c).** Plots show true values on the $x-$axis and the estimated values on the $y-$axis. For each plot, 80% HPD intervals, which covers the truth, are shown in blue, and 80% intervals, which do not cover the truth, are shown in red. The coverage percentages are shown in the top left of each plot.

##### S5.4 Simulated dataset using Visitor SIR model with five locations

For each set of visit depart and visit return rates described in Table S4 (Case(a) in Fig. 2, main text), we simulated 116 transmission trees from the Visitor SIR model with 5 locations. Then, we conducted a Bayesian coverage experiment using the same simulating model under Visitor SIR. Figure S14 shows the coverage levels across different parameters when fit to the the Visitor SIR model with four locations for 116 transmission trees simulated under the same model. In summary, most of the parameters across different movement scenarios, as described in Table S4, have coverage within the expectation. One exception is the the coverage for $R_0$ in Figure S14(c) that seems lower. However, this may be caused by some simulated trees being already past the exponential phase of the outbreak. Next, comparing with the migration models, as seen in Figures S16−S17, the coverage for the within-location and between-location infection rates are always low in both migration models (Cladogenetic Migration SIR and Simple Migration SIR).

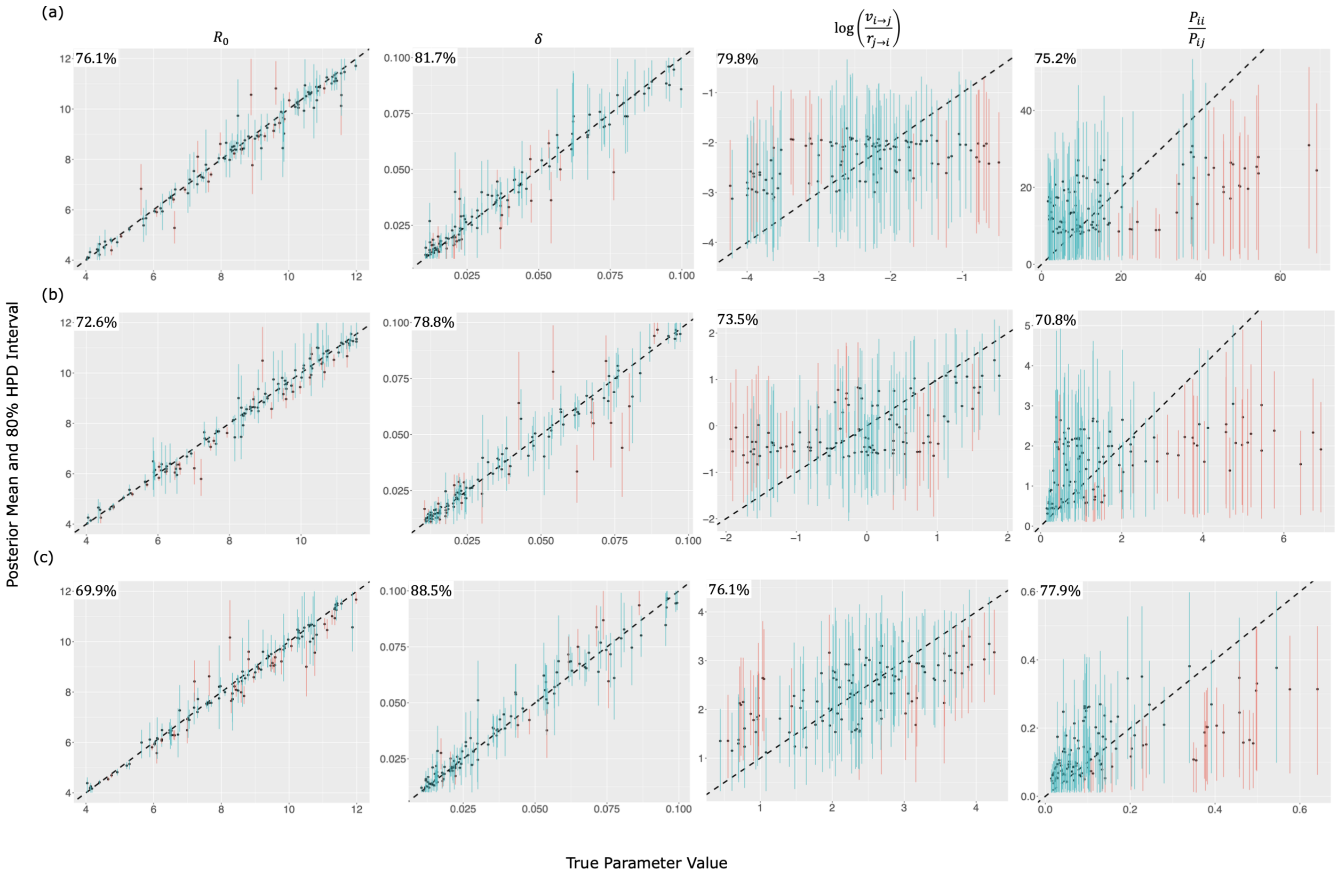


**Figure S14: Quality of parameter estimation and Bayesian coverage of the Visitor SIR model parameters in simulations with five locations and under all scenarios described in Table S4, namely (a) depart rate < return rate (top panel), (b) depart rate = return rate (middle panel), (c) depart rate > return rate (bottom panel).** Plots show true parameter values on the $x-$axis and estimated values on the $y-$axis. Parameter names are shown in the top of the upper plots, and the 80% HPD intervals, which cover the true values, are shown in blue, and the intervals, which do not cover the true values, are shown in red. The coverage percentages are shown in the top left of each plot.

**Table S4: Table showing priors for each estimated parameters with their definitions for each different model across three simulation scenarios under the Visitor SIR model.**

| Notation | Visitor SIR | Clado Migration SIR | Simple Migration SIR | Description |
|---|---|---|---|---|
| | Priors | | | |
| **(a) 5 locations, depart rate < return rate** | | | | |
| $R_0$ | U(4,12) | U(4,12) | U(4,12) | basic reproduction number |
| $\gamma$ | U(0.1,1) | U(0.1,1) | U(0.1,1) | recovery rate |
| $\delta$ | LogU(0.01,0.1) | LogU(0.01,0.1) | LogU(0.01,0.1) | sampling rate |
| $v_{i \to j}, \forall i, j$ | $\text{LogU}\left(\frac{2(0.01)}{4}, \frac{2(0.1)}{4}\right)$ | – | – | depart rate |
| $r_{j \to i}, \forall i, j$ | $\text{LogU}\left(\frac{2(0.1)}{4}, \frac{2(1.0)}{4}\right)$ | – | – | return rate |
| $m_{i \to j}, \forall i, j$ | – | $\text{LogU}\left(\frac{2(0.01)}{4}, \frac{2(0.1)}{4}\right)$ | $\text{LogU}\left(\frac{2(0.01)}{4}, \frac{2(0.1)}{4}\right)$ | migration rate |
| $p$ | – | U(0,1) | U(0,1) | Cladogenetic probability of observing no change in state |
| **(b) 5 locations, depart rate = return rate** | | | | |
| $R_0$ | U(4,12) | U(4,12) | U(4,12) | basic reproduction number |
| $\gamma$ | U(0.1,1) | U(0.1,1) | U(0.1,1) | recovery rate |
| $\delta$ | LogU(0.01,0.1) | LogU(0.01,0.1) | LogU(0.01,0.1) | sampling rate |
| $v_{i \to j}, \forall i, j$ | $\text{LogU}\left(\frac{2(0.1)}{4}, \frac{2(1.0)}{4}\right)$ | – | – | depart rate |
| $r_{j \to i}, \forall i, j$ | $\text{LogU}\left(\frac{2(0.1)}{4}, \frac{2(1.0)}{4}\right)$ | – | – | return rate |
| $m_{i \to j}, \forall i, j$ | – | $\text{LogU}\left(\frac{2(0.1)}{4}, \frac{2(1.0)}{4}\right)$ | $\text{LogU}\left(\frac{2(0.1)}{4}, \frac{2(1.0)}{4}\right)$ | migration rate |
| $p$ | – | U(0,1) | U(0,1) | Cladogenetic probability of observing no change in state |
| **(c) 5 locations, depart rate > return rate** | | | | |
| $R_0$ | U(4,12) | U(4,12) | U(4,12) | basic reproduction number |
| $\gamma$ | U(0.1,1) | U(0.1,1) | U(0.1,1) | recovery rate |
| $\delta$ | LogU(0.01,0.1) | LogU(0.01,0.1) | LogU(0.01,0.1) | sampling rate |
| $v_{i \to j}, \forall i, j$ | $\text{LogU}\left(\frac{2(0.1)}{4}, \frac{2(1.0)}{4}\right)$ | – | – | depart rate |
| $r_{j \to i}, \forall i, j$ | $\text{LogU}\left(\frac{2(0.01)}{4}, \frac{2(0.1)}{4}\right)$ | – | – | return rate |
| $m_{i \to j}, \forall i, j$ | – | $\text{LogU}\left(\frac{2(0.1)}{4}, \frac{2(1.0)}{4}\right)$ | $\text{LogU}\left(\frac{2(0.1)}{4}, \frac{2(1.0)}{4}\right)$ | migration rate |
| $p$ | – | U(0,1) | U(0,1) | Cladogenetic probability of observing no change in state |

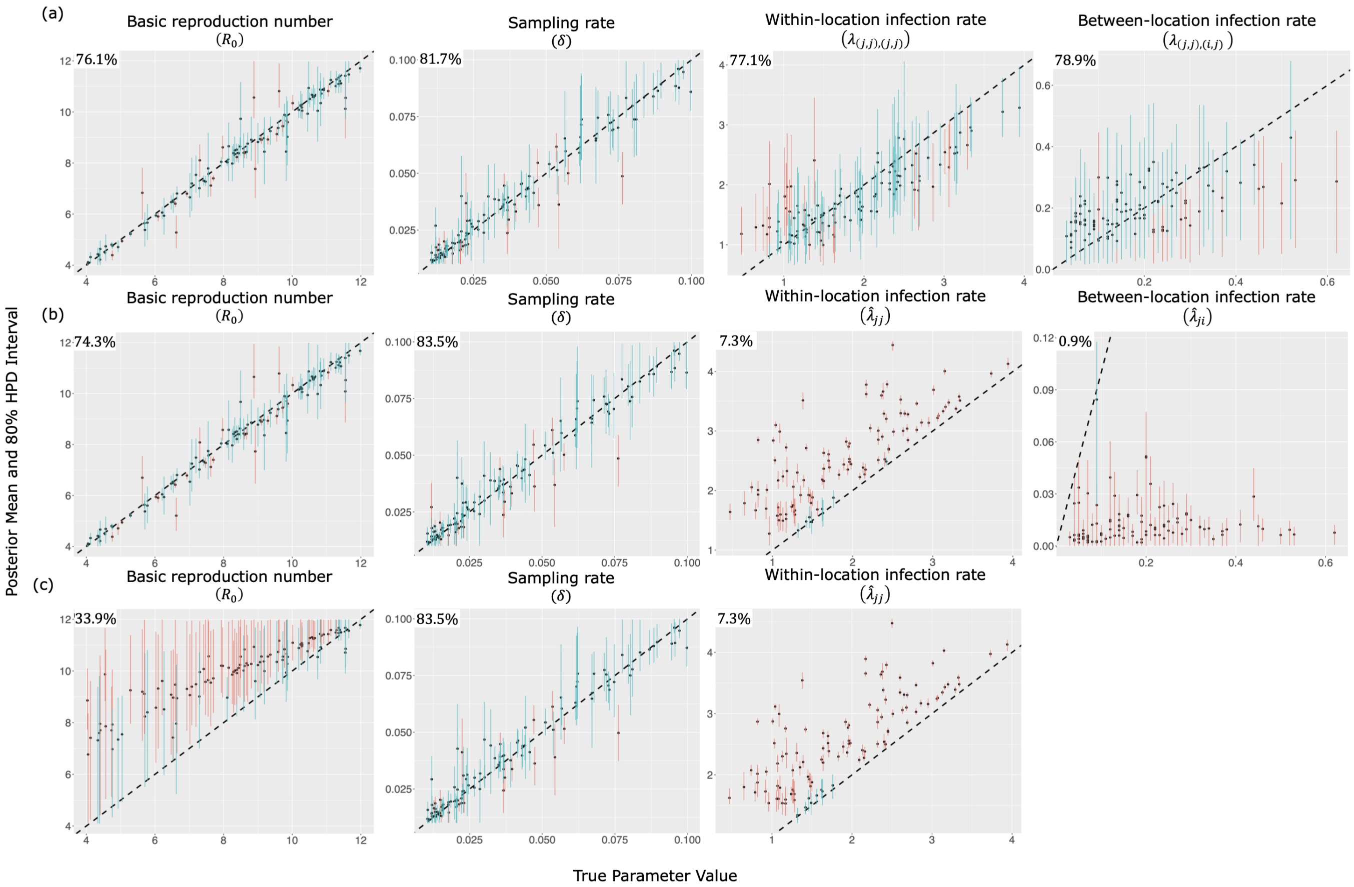


**Figure S15: Quality of parameter estimation and coverage across the inference models, namely (a) the Visitor SIR, (b) Cladogenetic Migration SIR, and (c) Simple Migration SIR models where the simulation model (the true model) is simulated under the same Visitor SIR model with 5 locations as the Visitor SIR model used for inference, as described in Table. S4(a).** Plots show true values on the $x-$axis and the estimated values on the $y-$axis. For each plot, 80% HPD intervals, which covers the truth, are shown in blue, and 80% intervals, which do not cover the truth, are shown in red. The coverage percentages are shown in the top left of each plot.

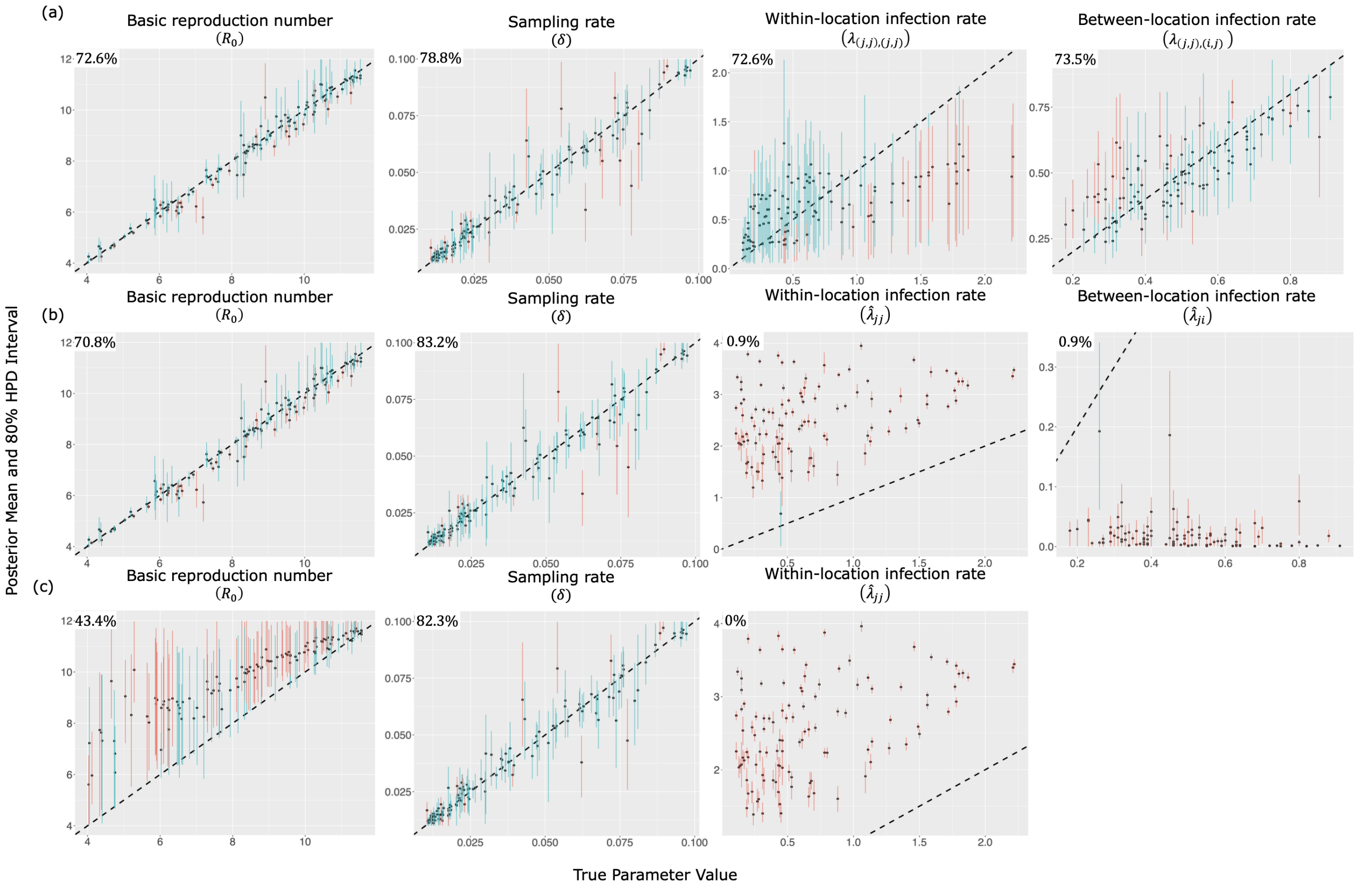


**Figure S16: Quality of parameter estimation and coverage across the inference models, namely (a) the Visitor SIR, (b) Cladogenetic Migration SIR, and (c) Simple Migration SIR models where the simulation model (the true model) is simulated under the same Visitor SIR model with 5 locations as the Visitor SIR model used for inference, as described in Table. S4(b).** Plots show true values on the $x-$axis and the estimated values on the $y-$axis. For each plot, 80% HPD intervals, which covers the truth, are shown in blue, and 80% intervals, which do not cover the truth, are shown in red. The coverage percentages are shown in the top left of each plot.

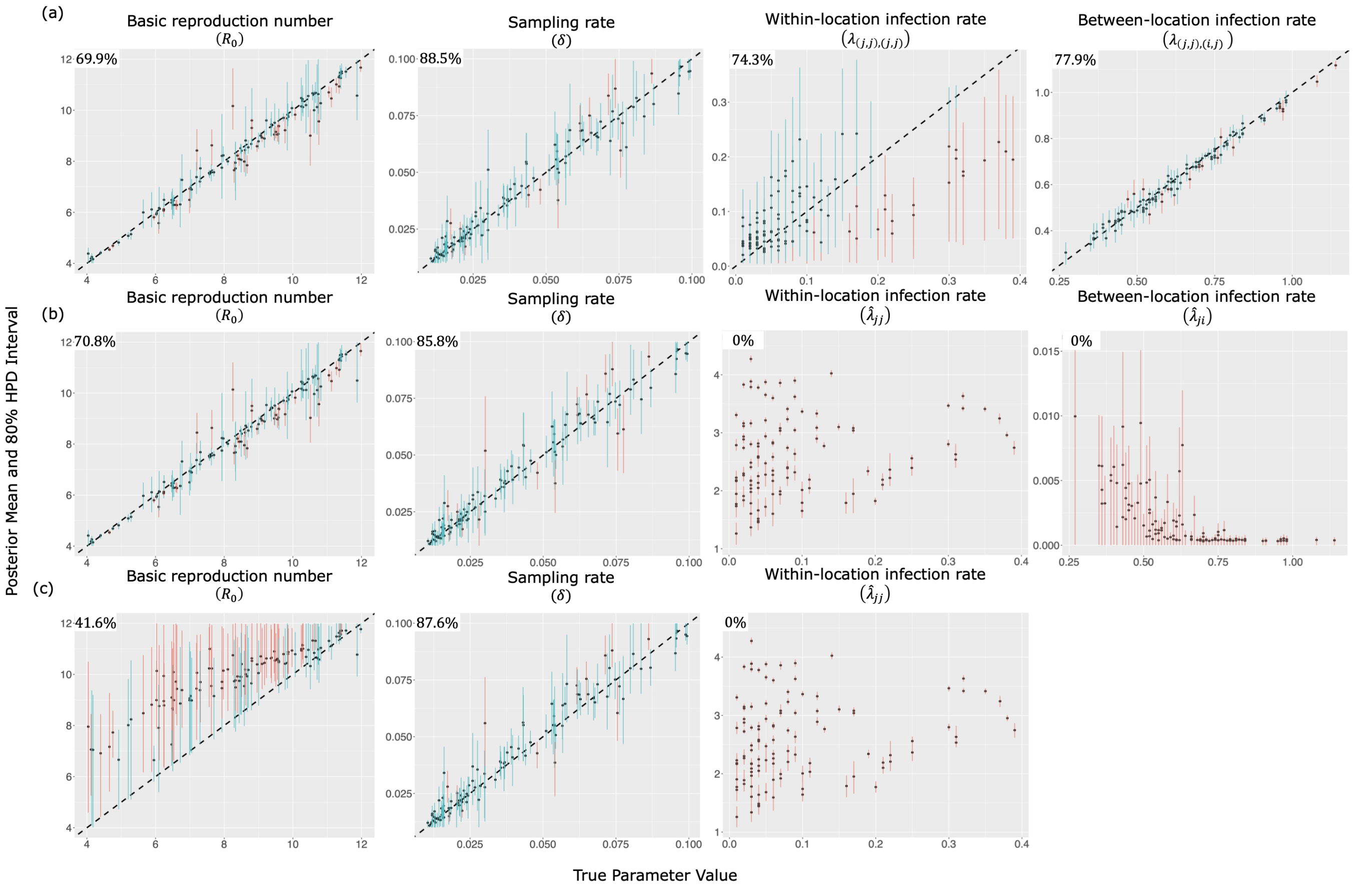


**Figure S17: Quality of parameter estimation and coverage across the inference models, namely (a) the Visitor SIR, (b) Cladogenetic Migration SIR, and (c) Simple Migration SIR models where the simulation model (the true model) is simulated under the same Visitor SIR model with 5 locations as the Visitor SIR model used for inference, as described in Table. S4(c).** Plots show true values on the $x-$axis and the estimated values on the $y-$axis. For each plot, 80% HPD intervals, which covers the truth, are shown in blue, and 80% intervals, which do not cover the truth, are shown in red. The coverage percentages are shown in the top left of each plot.

### S6 Transforming visitor state space for empirical analysis

We assume the following labels for the listed countries: 0 = Hubei, 1 = France, 2 = Germany, 3 = Italy, 4 = Other European nations. Thus, our empirical state space is. $S_e = \{0, 1, 2, 3, 4\}$. These are the actual (current) locations where infected individuals are sampled. However, the Visitor SIR model requires a compound state, consisting of both home and current locations. Therefore, we apply the following transformation, as shown in Eq. (23) to assign each current location with ambiguous home locations. We say a home location is ambiguous if any of these 5 locations above can be a home location for a particular infected individual in his current location.

| home\current | 0 | 1 | 2 | 3 | 4 |
|---|---|---|---|---|---|
| 0 | 0 | 1 | 2 | 3 | 4 |
| 1 | 5 | 6 | 7 | 8 | 9 |
| 2 | $A$ | $B$ | $C$ | $D$ | $E$ |
| 3 | $F$ | $G$ | $H$ | $I$ | $J$ |
| 4 | $K$ | $L$ | $M$ | $N$ | $O$ |

(23)

Each row in the matrix above corresponds to a home location and each column corresponds to a known current location. Therefore, under this transformation, our original state space $S_e$ becomes the transformed state space $\hat{S}_e$ under the visitor SIR model where,

$$\hat{S}_e = \{05AFK, 16BGL, 27CHM, 38DIN, 49EJO\} \tag{24}$$

### S7 Countries with missing trips and nights data

**Table S5: List of country pairs with missing travel information on number of nights spent and total number of trips.** Note that we have excluded pairs between countries that belong to Other European nations.

| From location | To location |
|---|---|
| Switzerland | China |
| Czech Republic | China |
| Germany | Finland |
| Germany | Iceland |
| Germany | Luxembourg |
| Germany | Slovakia |
| Denmark | China |
| Finland | China |
| France | Iceland |
| France | Slovakia |
| Ireland | China |
| Iceland | China |
| Iceland | Germany |
| Iceland | France |
| Iceland | Italy |
| Italy | Belgium |
| Italy | China |
| Italy | Denmark |
| Italy | Finland |
| Italy | Ireland |
| Italy | Iceland |
| Italy | Luxembourg |
| Italy | Norway |
| Italy | Poland |
| Italy | Portugal |
| Italy | Sweden |
| Italy | Slovakia |
| Norway | China |
| Norway | Germany |
| Norway | France |
| Norway | Italy |
| Portugal | China |
| Slovakia | China |
| Slovakia | France |
| United Kingdom | China |
| United Kingdom | Germany |
| United Kingdom | France |
| United Kingdom | Italy |

### S8 Ancestral state reconstruction under the Visitor SIR model

#### S8.1 Labeled by home locations

Here we labeled branches according to each individual's home location (Fig. S18).

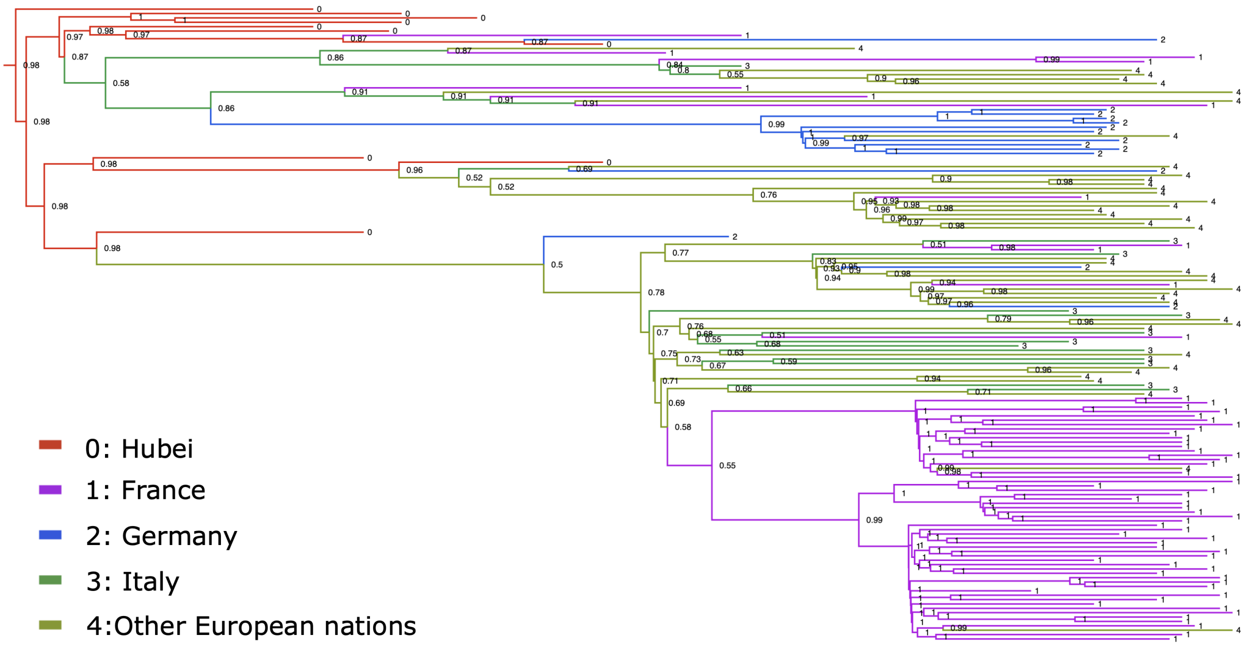


**Figure S18: The estimated ancestral state reconstruction according to individual's home location under our Visitor SIR mode with the most probable posterior support shown at each node.** The ancestral states are mapped onto the tree from Nadeau et al. (2021). The different colors represent individual's home location (at the start of each branch).

#### S8.2 Labeled by current locations

Here we labeled branches according to each individual's current location (Fig. S19).

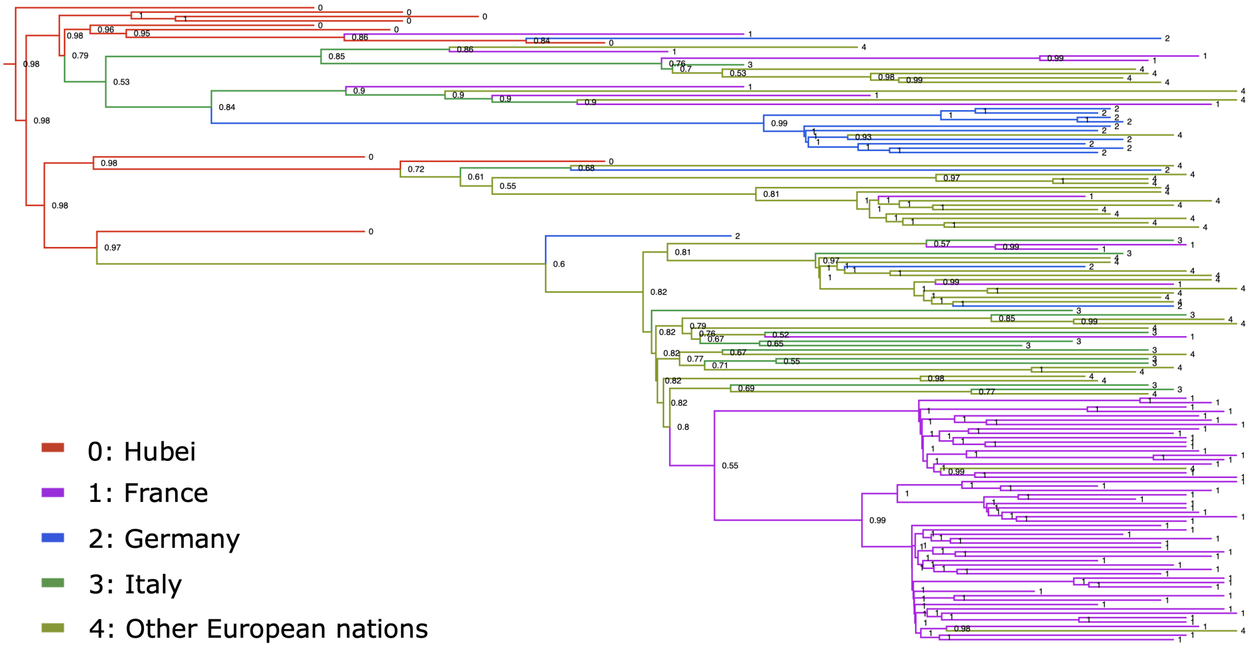


**Figure S19: The estimated ancestral state reconstruction according to individual's current location under our Visitor SIR model with the most probable posterior support shown at each node.** The ancestral states are mapped onto the tree from Nadeau et al. (2021). The different colors represent individual's current location (at the end of each branch).

##### S8.3 Labeled by matched locations

Here we labeled branches according to whether an individual is currently visiting a location or not visiting. (Fig. S19). Using the empirical tree from Nadeau et al. (2021) we found one lineage (indicated by the red branch) that corresponds to a visitor following an infection.

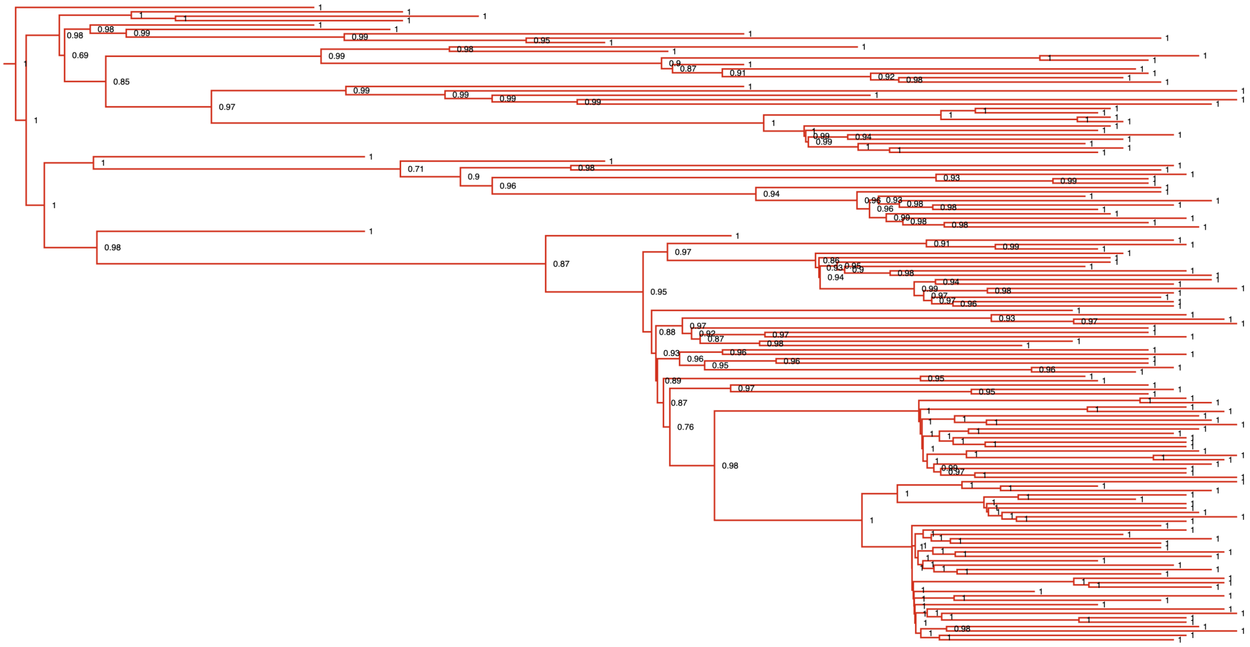

**Figure S20: The estimated ancestral state reconstruction under our Visitor SIR mode with the most probable posterior support shown at each node.** The ancestral states are mapped onto the tree from Nadeau et al. (2021). Red color represents cases the home and away locations of an individual lineage match.
